# Plasma sgp130 is an independent predictor of non-alcoholic fatty liver disease severity

**DOI:** 10.1101/2022.01.10.22268968

**Authors:** Aysim Gunes, Laurent Bilodeau, Catherine Huet, Assia Belblidia, Cindy Baldwin, Jeanne-Marie Giard, Laurent Biertho, Annie Lafortune, Christian Yves Couture, Bich N Nguyen, Eithan Galun, Chantal Bémeur, Marc Bilodeau, Mathieu Laplante, An Tang, May Faraj, Jennifer L. Estall

**Affiliations:** Institut de recherches cliniques de Montréal (IRCM), Montréal, Québec, Canada; Division of Experimental Medicine, McGill University, Montreal, Quebec, Canada; Montreal Diabetes Research Centre, Montreal, Quebec, Canada; Département de radiologie, Centre hospitalier de l’Université de Montréal (CHUM), Montréal, Québec, Canada; Département d’hépatologie, Centre hospitalier de l’Université de Montréal (CHUM), Département de médecine, Université de Montréal, Centre de recherche du CHUM, Montréal, Québec, Canada; Centre de recherche de l’Institut universitaire de cardiologie et de pneumologie de Québec, Université Laval, Québec, Canada; Département de chirurgie, Faculté de médecine, Université Laval, Québec, Canada; Département de biologie moléculaire, biochimie médicale et pathologie, Université Laval, Québec, Canada; Département de pathologie et biologie cellulaire, Université de Montréal, Montréal, Québec, Canada; Goldyne Savad Institute of Gene Therapy, Hadassah Hebrew University Hospital, Jerusalem, Israel; Département de nutrition, Université de Montréal, Montréal, Québec, Canada; Labo HépatoNeuro, Centre de recherche du CHUM, Montréal, Québec, Canada; Centre de recherche sur le cancer de l’Université Laval, Université Laval, Québec, QC, Canada

**Keywords:** IL-6 transsignaling, NAFLD, NASH, liver fibrosis

## Abstract

**Background:** Interleukin-6 (IL-6) plays important and dynamic roles in inflammation associated with fatty liver disease over all stages, from simple steatosis to steatohepatitis, cirrhosis and cancer. IL-6 signals locally, but also circulates with multiple co-factors that control paracrine and endocrine signaling. As inflammation is a main driver of liver fibrosis, we investigated relationships between circulating components of the interleukin-6 signaling pathway (IL-6, sIL-6R and sgp130) and liver pathology in subjects with metabolically associated fatty liver disease (MAFLD) or steatohepatitis (MASH).

**Methods:** Predictive performances of plasma IL-6, sIL-6R and sgp130 were investigated in two independent cohorts: 1) patients with biopsy-confirmed MASH (*n*=49), where magnetic resonance spectroscopy (MRS), imaging (MRI) and elastography (MRE) assessed liver fat, volume and stiffness; and 2) patients with morbid obesity (*n*=245) undergoing bariatric surgery where histological staging of steatosis, activity, and fibrosis determined MASH severity. Correlations were evaluated between IL-6, sIL-6R and sgp130 and anthropomorphic characteristics, plasma markers of metabolic disease or liver pathology.

**Results:** In patients with MASH, plasma IL-6 and sgp130 strongly correlated with liver stiffness, which for sgp130 was independent of age, sex, BMI, diabetes, hyperlipidemia, hypertension or history of HCC. Plasma sgp130 was the strongest predictor of liver stiffness compared to common predictors and risk scores. Plasma sIL-6R correlated with liver volume independent of age, sex, and BMI. In patients with morbid obesity, circulating sgp130 correlated with advanced liver fibrosis.

**Conclusion:** Levels of circulating sgp130 can predict progressing MASH and may be used alone or in combination with other predictors as a non-invasive measure of liver disease severity.

**GRAPHICAL ABSTRACT:** 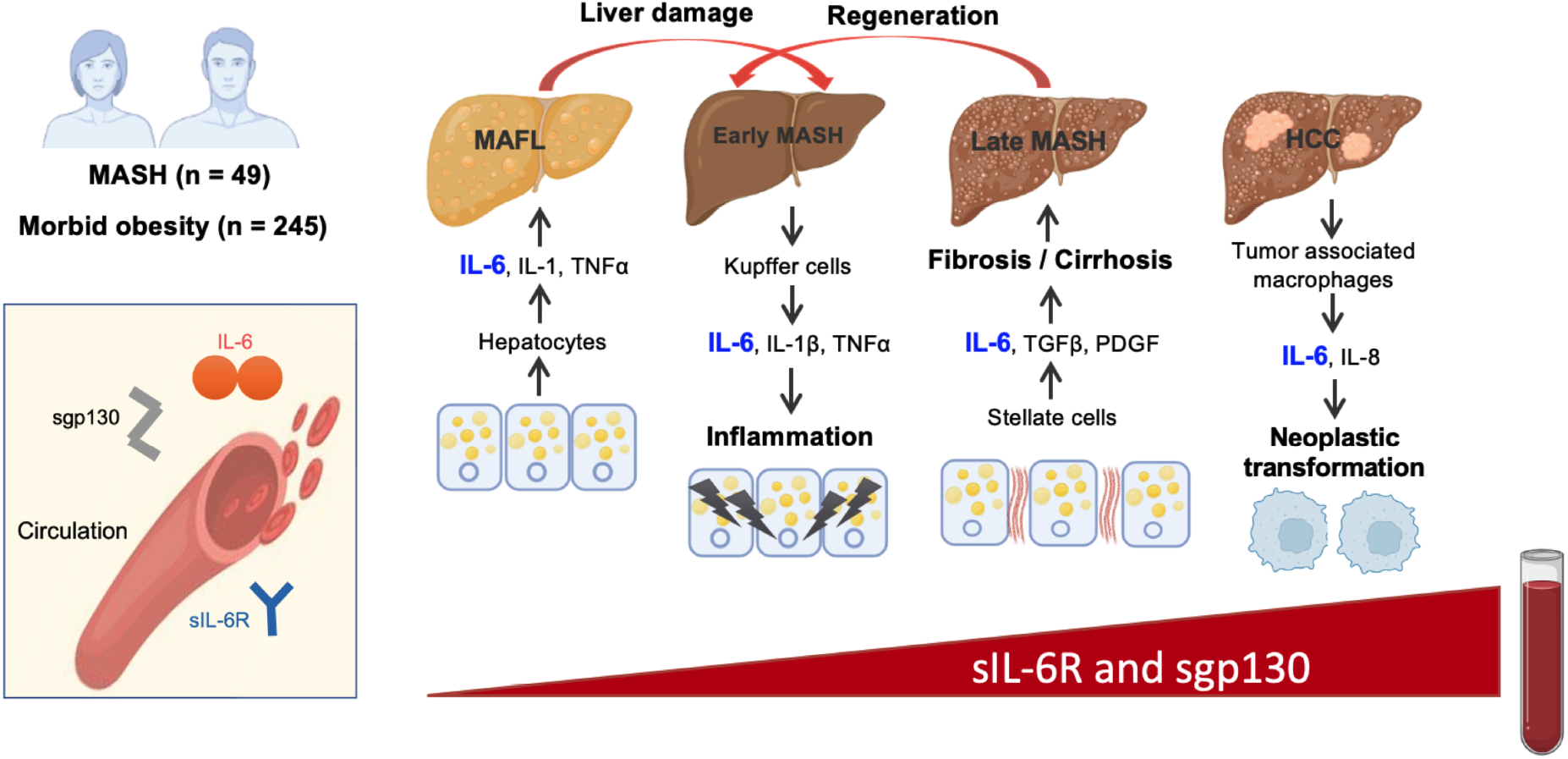

## INTRODUCTION

Nonalcoholic fatty liver disease (NAFLD) is rapidly increasing worldwide, with global prevalence of disease reaching approximately 25% (1–3). NAFLD is a multistep, progressive disorder beginning with simple steatosis that can evolve to non-alcoholic steatohepatitis (NASH), characterized by hepatocellular ballooning, lobular inflammation, and a range of fibrosis. Simple steatosis is relatively benign, while NASH can progress to cirrhosis and/or hepatocellular carcinoma (HCC) (4, 5). Steatohepatitis is now considered the second most common indication for liver transplantation and HCC in Canada (6).

Fatty liver disease stemming from metabolic disorders has garnered the unique sub-designation of MAFLD or MASH, due to its distinct etiology and progression. The transition from simple steatosis to steatohepatitis is an important stage of fatty liver disease correlating strongly with poor prognosis (7). Liver fibrosis is a primary predictor of morbidity and mortality (8). Each advancing stage adversely affects the health and survival of patients, yet we do not yet know or understand all the factors that promote these steps. Our ability to identify these factors is limited by the asymptomatic nature of the simple steatosis to MASH transition, preventing early diagnosis and study of the disease.

Initial diagnoses often use liver risk scores calculated from age, body mass index (BMI), diabetes status, and plasma liver enzymes (alanine and aspartate aminotransferases, ALT and AST) to predict NAFLD progression and severity. However, recent evidence suggests that BMI poorly correlates with NAFLD severity (9) and as high as 80% of subjects with NAFLD and 19% of subjects with biopsy-proven NASH have normal liver enzyme levels (10–13). Thus, relying on obesity and increased liver enzymes to screen for NAFLD can lead to underdiagnosis and/or underestimation of liver damage. Liver inflammation plays a key role in the transition to steatohepatitis, yet currently, there are no non-invasive, sensitive indicators of the early inflammatory stages of NASH.

Interleukin-6 (IL-6) is an inflammatory cytokine closely associated with metabolic disease (14–18). It has both pro- and anti-inflammatory properties and may play a role in the progression of NAFLD to NASH (19, 20), cirrhosis (21) and liver cancer (22). IL-6 signaling involves either activation of a membrane-bound receptor (classical) or formation of a signaling complex with a soluble IL-6 receptor (sIL-6R) found in circulation (trans-signaling). Both types of signaling require the ligand/receptor complex to bind a co-receptor, glycoprotein 130 (gp130), on the surface of cells. Circulating sIL-6R, shed from receptor-expressing cells, can dock to gp130 on distant cells, allowing IL-6 (trans)signaling in tissues that do not express the IL-6 receptor. A soluble, secreted form of the co-receptor (sgp130) also circulates and can inhibit trans-signaling by sequestering the IL-6/sIL-6R complex (23). Based on expression patterns, hepatocytes and hepatic stellate cells may be a rich source of sIL-6R and sgp130, respectively, suggesting the liver as a potential major player in IL-6 trans-signaling (24, 25). There is evidence linking increased IL-6 trans-signaling to other metabolic diseases (17, 26, 27), as well as to alcohol- and infection-induced chronic liver disease (28).

Given the involvement of IL-6 signaling at all stages of NAFLD and its link to metabolic disease, we hypothesized that circulating mediators of the IL-6 trans-signaling pathway may predict fatty liver disease severity in populations with MASH and/or morbid obesity. In this study, we investigated relationships between circulating IL-6, sIL-6R and sgp130, and liver pathology associated with metabolic syndrome in two human cohorts.

## RESULTS

### Characteristics of patients with MASH

Anthropometric, metabolic, and clinical characteristics of our first cohort (28 women and 21 men) are presented in **Table 1**. Subjects chosen met the current criteria of MAFLD/MASH, which requires either obesity, diagnosis of type 2 diabetes mellitus, or evidence of metabolic dysregulation (9) and elimination of other possible NAFLD causes. Among the population: 44.9% of subjects had diabetes; 49.0% had obesity; 28.6% had hyperlipidemia; 44.9% had hypertension; and, 40.8% consumed alcohol in moderation. Hepatic histological evaluation of patients revealed that 75.0% had stage 2 steatosis, 70.0% had an activity score of 2, and 87.5% had stage 1-2 fibrosis at the time of the biopsy **(Table 1)**, suggesting that most were in the early stages of MASH. MRI/MRE measures of liver fat, volume and stiffness were affected by comorbidities in patients with MASH. Liver fat fraction was lower in patients with a previous history of HCC (p = 0.004); liver volume was significantly higher in patients with diabetes (p = 0.004) and/or obesity (p = 0.004); and liver stiffness was higher in patients with diabetes (p = 0.005), obesity (p = 0.003), hypertension (p = 0.022) and/or a previous history of HCC (p = 0.003) **(Supplementary Figure 2A-R)**. As expected in MASH, plasma globulin, ALT, GGT, glycemia and HbA1c levels were higher than normal ranges in the Canadian population (29) (**Table 1**). In line with expected sex differences, blood platelet count was lower (p = 0.036), and plasma AST (p = 0.034) and GGT were higher (p = 0.017), in men compared to women (29).

**Table 1.**
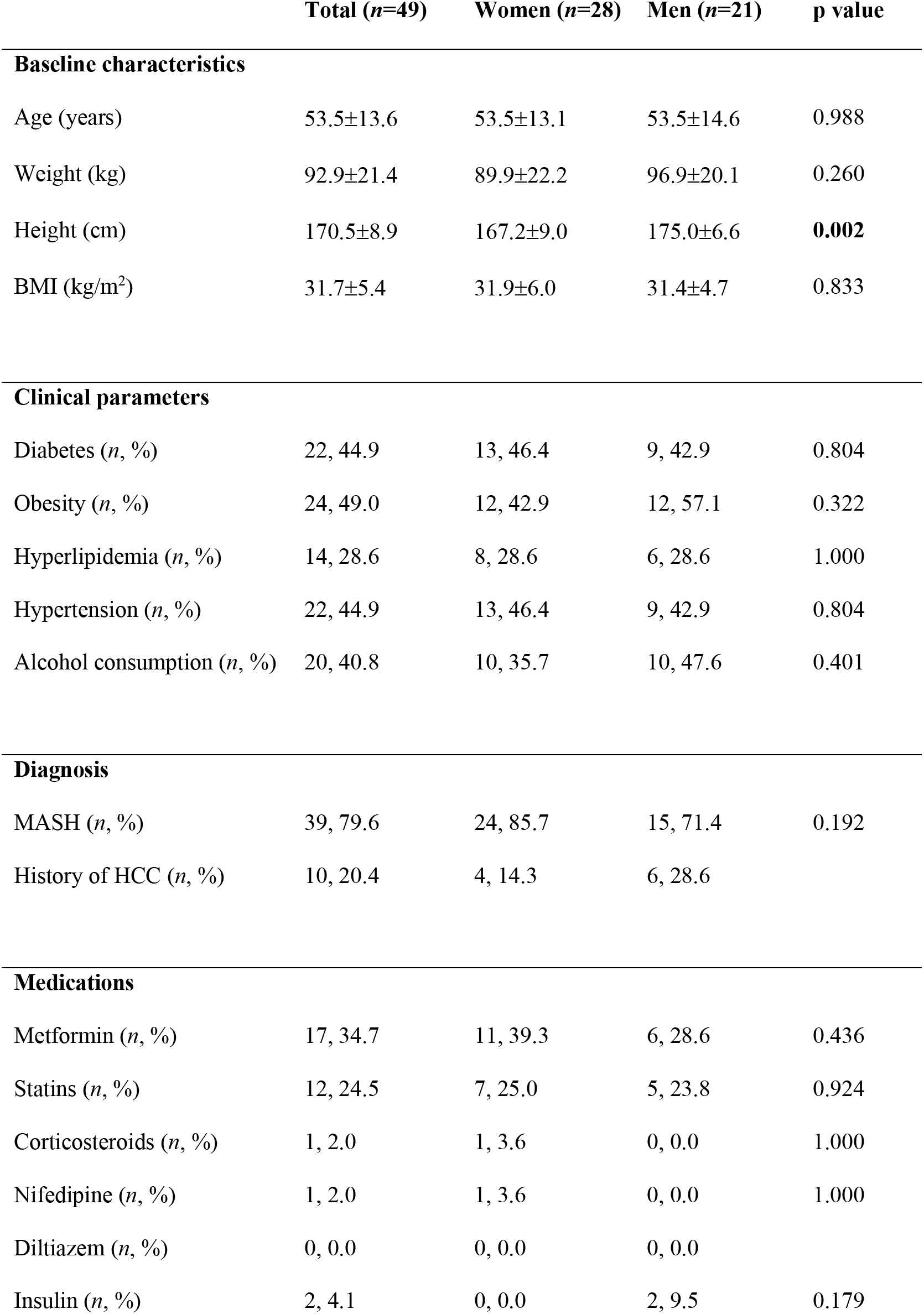
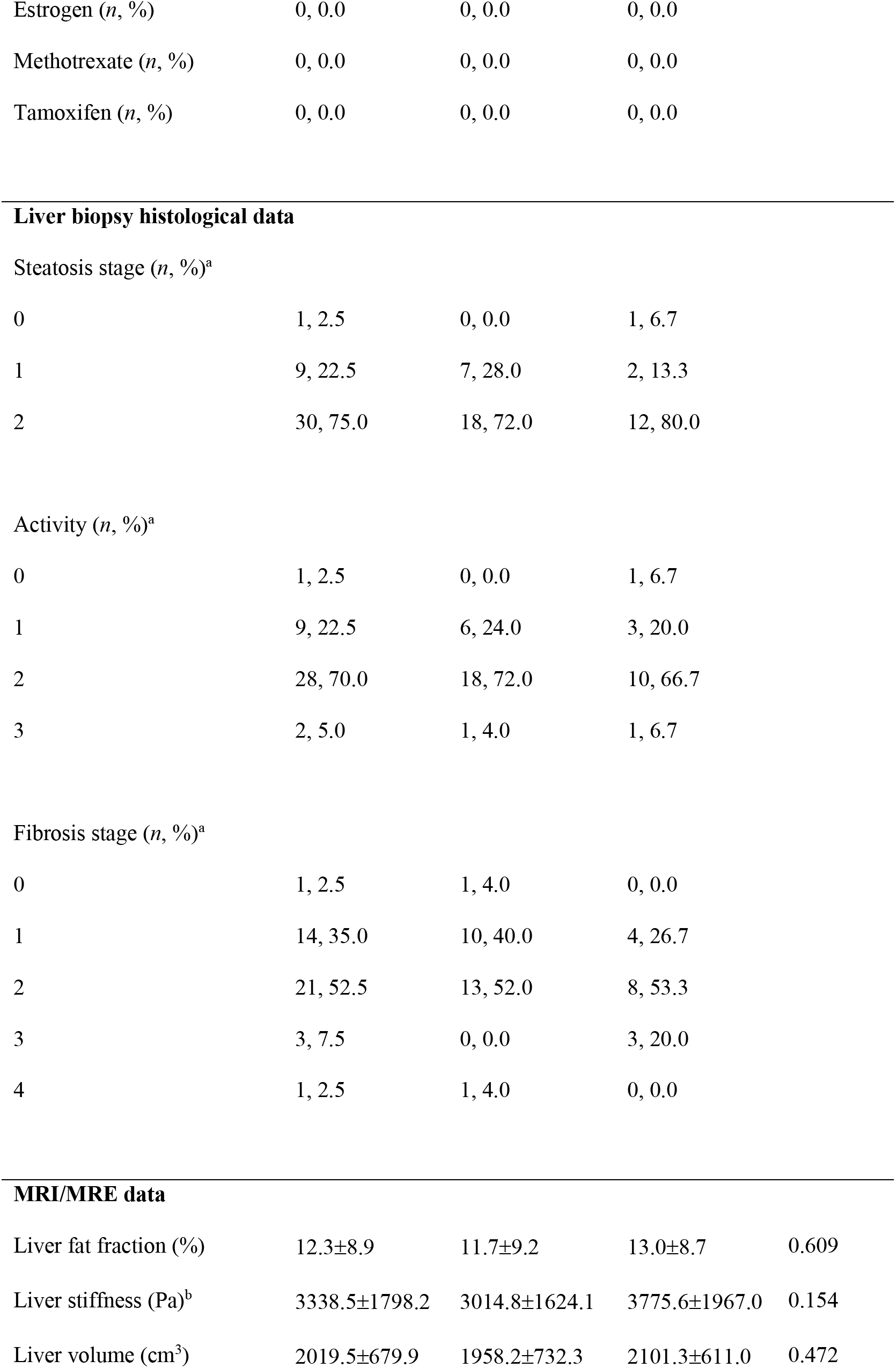
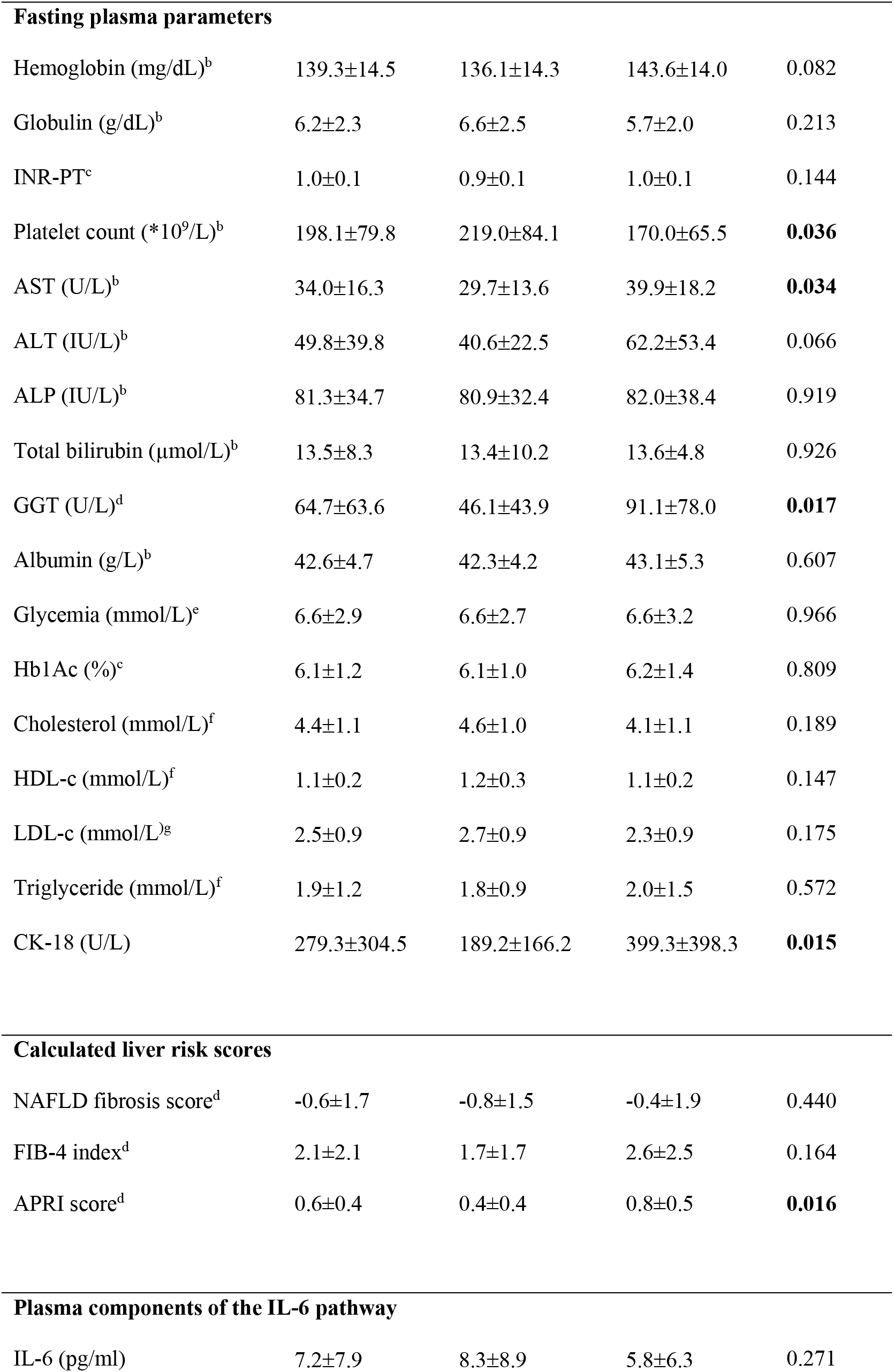
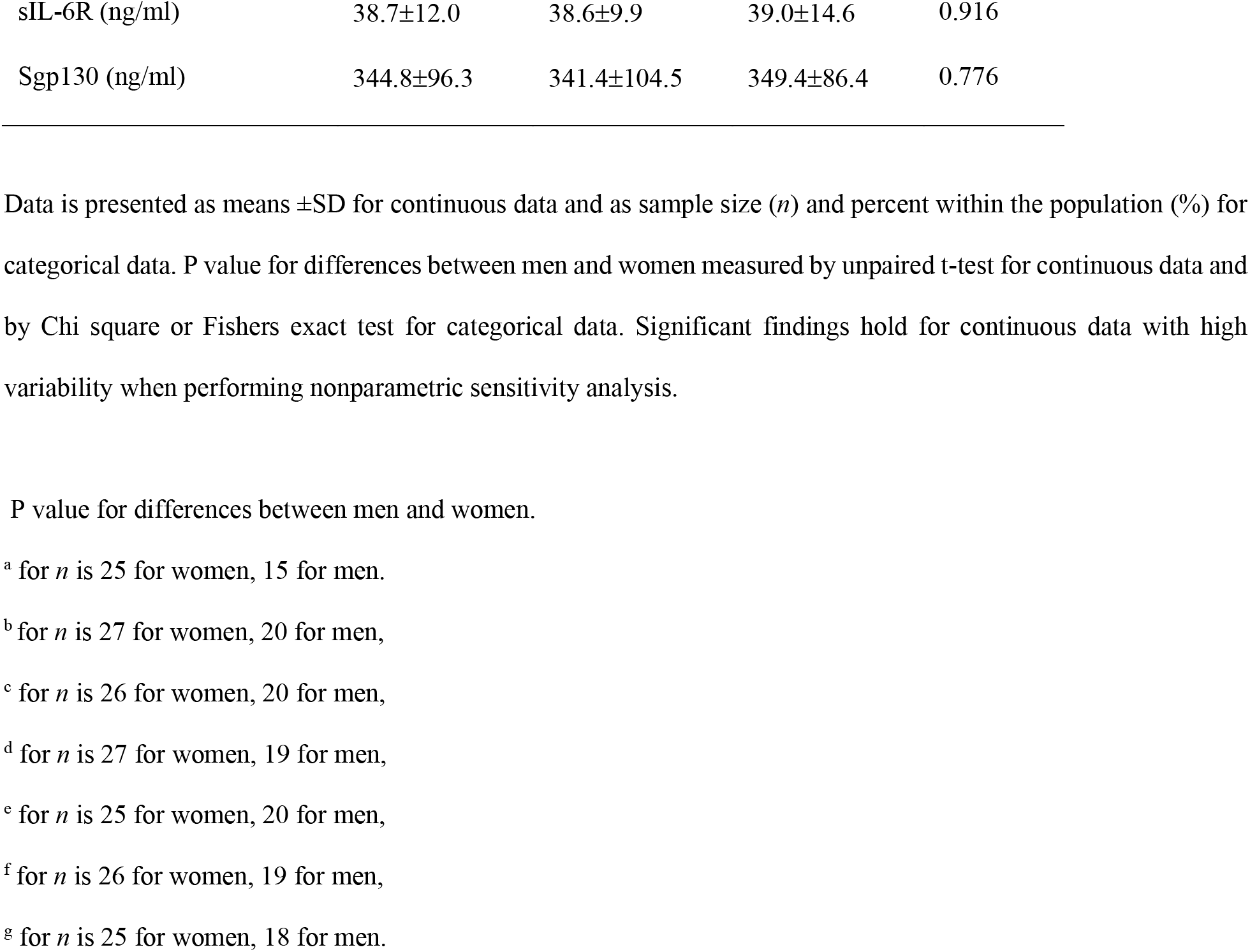
Anthropometric, metabolic and clinical characteristics of patients with MASH.

|  | **Total (n=49)** | **Women (n=28)** | **Men (n=21)** | **p value** |
| --- | --- | --- | --- | --- |
| **Baseline characteristics** |  |  |  |  |
| Age (years) | 53.5±13.6 | 53.5±13.1 | 53.5±14.6 | 0.988 |
| Weight (kg) | 92.9±21.4 | 89.9±22.2 | 96.9±20.1 | 0.260 |
| Height (cm) | 170.5±8.9 | 167.2±9.0 | 175.0±6.6 | **0.002** |
| BMI (kg/m<sup>2</sup>) | 31.7±5.4 | 31.9±6.0 | 31.4±4.7 | 0.833 |
| **Clinical parameters** |  |  |  |  |
| Diabetes (n, %) | 22, 44.9 | 13, 46.4 | 9, 42.9 | 0.804 |
| Obesity (n, %) | 24, 49.0 | 12, 42.9 | 12, 57.1 | 0.322 |
| Hyperlipidemia (n, %) | 14, 28.6 | 8, 28.6 | 6, 28.6 | 1.000 |
| Hypertension (n, %) | 22, 44.9 | 13, 46.4 | 9, 42.9 | 0.804 |
| Alcohol consumption (n, %) | 20, 40.8 | 10, 35.7 | 10, 47.6 | 0.401 |
| **Diagnosis** |  |  |  |  |
| MASH (n, %) | 39, 79.6 | 24, 85.7 | 15, 71.4 | 0.192 |
| History of HCC (n, %) | 10, 20.4 | 4, 14.3 | 6, 28.6 |  |
| **Medications** |  |  |  |  |
| Metformin (n, %) | 17, 34.7 | 11, 39.3 | 6, 28.6 | 0.436 |
| Statins (n, %) | 12, 24.5 | 7, 25.0 | 5, 23.8 | 0.924 |
| Corticosteroids (n, %) | 1, 2.0 | 1, 3.6 | 0, 0.0 | 1.000 |
| Nifedipine (n, %) | 1, 2.0 | 1, 3.6 | 0, 0.0 | 1.000 |
| Diltiazem (n, %) | 0, 0.0 | 0, 0.0 | 0, 0.0 |  |
| Insulin (n, %) | 2, 4.1 | 0, 0.0 | 2, 9.5 | 0.179 |

|  |  |  |  |
| --- | --- | --- | --- |
| Estrogen (*n*, %) | 0, 0.0 | 0, 0.0 | 0, 0.0 |
| Methotrexate (*n*, %) | 0, 0.0 | 0, 0.0 | 0, 0.0 |
| Tamoxifen (*n*, %) | 0, 0.0 | 0, 0.0 | 0, 0.0 |

Steatosis stage (*n*, %)<sup>a</sup>
|  |  |  |  |
| --- | --- | --- | --- |
| 0 | 1, 2.5 | 0, 0.0 | 1, 6.7 |
| 1 | 9, 22.5 | 7, 28.0 | 2, 13.3 |
| 2 | 30, 75.0 | 18, 72.0 | 12, 80.0 |

Activity (*n*, %)<sup>a</sup>
|  |  |  |  |
| --- | --- | --- | --- |
| 0 | 1, 2.5 | 0, 0.0 | 1, 6.7 |
| 1 | 9, 22.5 | 6, 24.0 | 3, 20.0 |
| 2 | 28, 70.0 | 18, 72.0 | 10, 66.7 |
| 3 | 2, 5.0 | 1, 4.0 | 1, 6.7 |

Fibrosis stage (*n*, %)<sup>a</sup>
|  |  |  |  |
| --- | --- | --- | --- |
| 0 | 1, 2.5 | 1, 4.0 | 0, 0.0 |
| 1 | 14, 35.0 | 10, 40.0 | 4, 26.7 |
| 2 | 21, 52.5 | 13, 52.0 | 8, 53.3 |
| 3 | 3, 7.5 | 0, 0.0 | 3, 20.0 |
| 4 | 1, 2.5 | 1, 4.0 | 0, 0.0 |
| MRI/MRE data |  |  |  |  |
| Liver fat fraction (%) | 12.3±8.9 | 11.7±9.2 | 13.0±8.7 | 0.609 |
| Liver stiffness (Pa)<sup>b</sup> | 3338.5±1798.2 | 3014.8±1624.1 | 3775.6±1967.0 | 0.154 |
| Liver volume (cm<sup>3</sup>) | 2019.5±679.9 | 1958.2±732.3 | 2101.3±611.0 | 0.472 |

| **Fasting plasma parameters** |  |  |  |  |
| --- | --- | --- | --- | --- |
| Hemoglobin (mg/dL)<sup>b</sup> | 139.3±14.5 | 136.1±14.3 | 143.6±14.0 | 0.082 |
| Globulin (g/dL)<sup>b</sup> | 6.2±2.3 | 6.6±2.5 | 5.7±2.0 | 0.213 |
| INR-PT<sup>c</sup> | 1.0±0.1 | 0.9±0.1 | 1.0±0.1 | 0.144 |
| Platelet count (*10<sup>9</sup>/L)<sup>b</sup> | 198.1±79.8 | 219.0±84.1 | 170.0±65.5 | **0.036** |
| AST (U/L)<sup>b</sup> | 34.0±16.3 | 29.7±13.6 | 39.9±18.2 | **0.034** |
| ALT (IU/L)<sup>b</sup> | 49.8±39.8 | 40.6±22.5 | 62.2±53.4 | 0.066 |
| ALP (IU/L)<sup>b</sup> | 81.3±34.7 | 80.9±32.4 | 82.0±38.4 | 0.919 |
| Total bilirubin (μmol/L)<sup>b</sup> | 13.5±8.3 | 13.4±10.2 | 13.6±4.8 | 0.926 |
| GGT (U/L)<sup>d</sup> | 64.7±63.6 | 46.1±43.9 | 91.1±78.0 | **0.017** |
| Albumin (g/L)<sup>b</sup> | 42.6±4.7 | 42.3±4.2 | 43.1±5.3 | 0.607 |
| Glycemia (mmol/L)<sup>c</sup> | 6.6±2.9 | 6.6±2.7 | 6.6±3.2 | 0.966 |
| Hb1Ac (%)<sup>c</sup> | 6.1±1.2 | 6.1±1.0 | 6.2±1.4 | 0.809 |
| Cholesterol (mmol/L)<sup>f</sup> | 4.4±1.1 | 4.6±1.0 | 4.1±1.1 | 0.189 |
| HDL-c (mmol/L)<sup>f</sup> | 1.1±0.2 | 1.2±0.3 | 1.1±0.2 | 0.147 |
| LDL-c (mmol/L)<sup>g</sup> | 2.5±0.9 | 2.7±0.9 | 2.3±0.9 | 0.175 |
| Triglyceride (mmol/L)<sup>f</sup> | 1.9±1.2 | 1.8±0.9 | 2.0±1.5 | 0.572 |
| CK-18 (U/L) | 279.3±304.5 | 189.2±166.2 | 399.3±398.3 | **0.015** |
| **Calculated liver risk scores** |  |  |  |  |
| NAFLD fibrosis score<sup>d</sup> | -0.6±1.7 | -0.8±1.5 | -0.4±1.9 | 0.440 |
| FIB-4 index<sup>d</sup> | 2.1±2.1 | 1.7±1.7 | 2.6±2.5 | 0.164 |
| APRI score<sup>d</sup> | 0.6±0.4 | 0.4±0.4 | 0.8±0.5 | **0.016** |
| **Plasma components of the IL-6 pathway** |  |  |  |  |
| IL-6 (pg/ml) | 7.2±7.9 | 8.3±8.9 | 5.8±6.3 | 0.271 |
| sIL-6R (ng/ml) | 38.7±12.0 | 38.6±9.9 | 39.0±14.6 | 0.916 |
| Sgp130 (ng/ml) | 344.8±96.3 | 341.4±104.5 | 349.4±86.4 | 0.776 |

Data is presented as means ±SD for continuous data and as sample size (*n*) and percent within the population (%) for categorical data. P value for differences between men and women measured by unpaired t-test for continuous data and by Chi square or Fishers exact test for categorical data. Significant findings hold for continuous data with high variability when performing nonparametric sensitivity analysis.
P value for differences between men and women.
<sup>a</sup> for *n* is 25 for women, 15 for men.
<sup>b</sup> for *n* is 27 for women, 20 for men,
<sup>c</sup> for *n* is 26 for women, 20 for men,
<sup>d</sup> for *n* is 27 for women, 19 for men,
<sup>e</sup> for *n* is 25 for women, 20 for men,
<sup>f</sup> for *n* is 26 for women, 19 for men,
<sup>g</sup> for *n* is 25 for women, 18 for men.

In healthy subjects, normal levels of plasma IL-6 are <3 pg/ml (29, 30) and average plasma concentrations are 35 ng/ml for sIL-6R and 217 ng/ml for sgp130 (30, 31). In this cohort, average plasma IL-6 and spg130 were above those reported for healthy subjects, while average levels of sIL-6R were close to normal **(Table 1**). Plasma IL-6 was significantly higher in MASH patients with diabetes (p = 0.029), hypertension (p = 0.005) or previous history of HCC (p = 0.025) compared to MASH patients without these co-morbidities (**Figure 1A, 1J, and 1P**). Plasma IL-6 also correlated positively with BMI (r = 0.34, p = 0.018, **Supplementary Table 1)** as previously reported in literature (15, 16). Plasma sIL-6R was higher in patients with diabetes (p = 0.007) **(Figure 1B)** while plasma sgp130 was higher in patients with hypertension (p = 0.034) or history of HCC (p < 0.0001) **(Figure 1L and 1R).**

**Figure 1.**
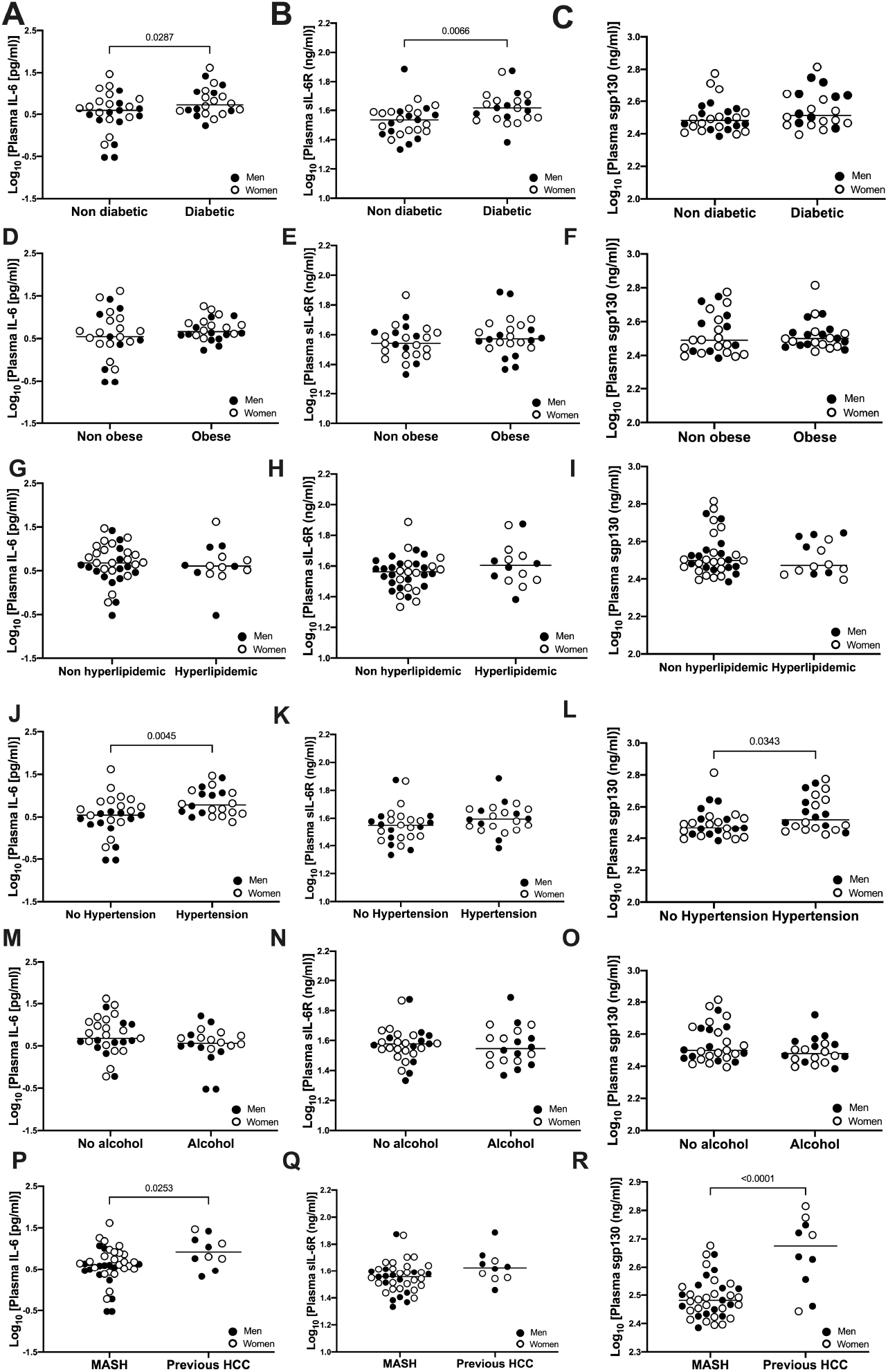
Plasma IL-6, sIL-6R and sgp130 levels in patients with MASH with or without diabetes (A-C), obesity (D-F), hyperlipidemia (H-I), hypertension (J-L), alcohol consumption (M-O) and previous history of HCC (P-R). Data is presented as the distribution around the mean. Analysis was conducted using unpaired t test. *n*=28 women (open circles) and *n*=21 men (closed circles).

### Plasma sgp130 predicts the severity of liver stiffness independent of age, sex, adiposity, and comorbidities in patients with MASH

Liver volume and liver stiffness (resistance of the tissue to deformation) are both accurately measured by MRI/MRE over the entire liver (32) and measurements of liver stiffness by MRI/MRE have good prognostic value to predict liver disease severity (33–35). Increased liver volume is influenced by several factors including fat content, inflammation, and/or edema, while increased liver stiffness is more indicative of increased inflammation and/or fibrosis (36). Of the three components of the IL-6 pathway, plasma sgp130 negatively correlated with liver fat fraction (r = −0.31, p = 0.031) (**Figure 2C)**, while plasma sIL-6R positively correlated with liver volume (r = 0.36, p = 0.011) (**Figure 2E)**. Both plasma IL-6 (r = 0.43, p = 0.002) and plasma sgp130 positively correlated with liver stiffness **(Figure 2G and 2I)**, with plasma sgp130 showing the strongest association (r = 0.77, p<0.0001).

**Figure 2.**
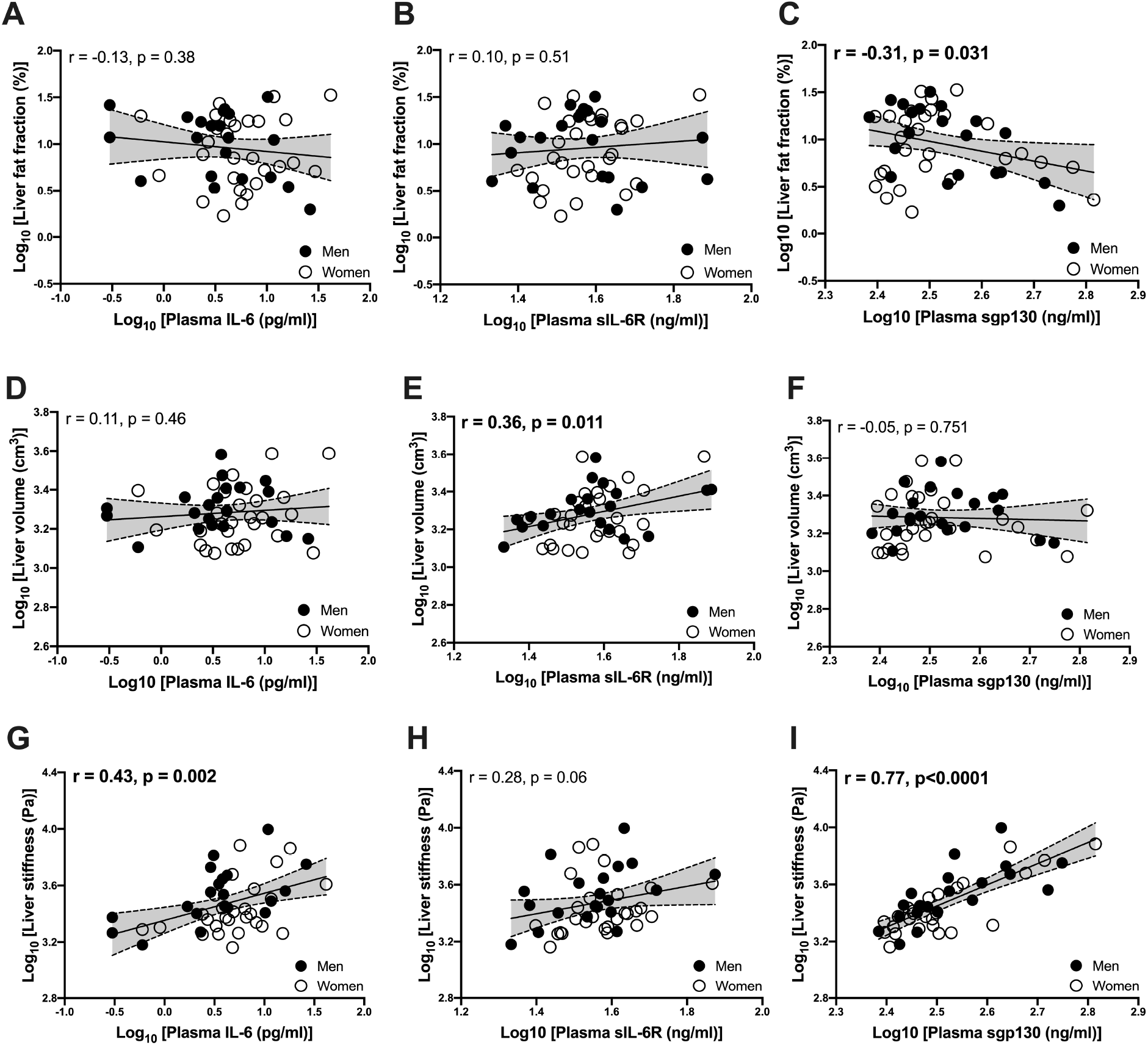
Pearson correlations between plasma IL-6, sIL-6R and sgp130 with MRE/MRI measures of liver fat fraction (A, B, C), liver volume (D, E, F) and liver stiffness (G, H, I) in patients with MASH. For liver fat fraction and liver volume, *n*=28 women (open circles) and *n*=21 men (closed circles). For liver stiffness, *n*=27 women (open circles) and *n*=20 men (closed circles).

Inclusion of subjects with a previous history of HCC allowed broader representation of liver disease severity within our cohort. While these subjects had no detectable cancer at the time of blood sampling, they had expectedly higher levels of liver stiffness (5416.5 ± 2517.9 Pa) and lower liver fat (4.6 ± 2.8 %). Concerned that this variable could influence the associations, we performed similar analysis while excluding subjects with history of HCC. In this subset (n = 39), the correlation between plasma sIL-6R and liver volume was strengthened (r = 0.457, p = 0.004) and the association between sgp130 and liver stiffness remained robust (r = 0.692, p<0.0001). The relationship between plasma IL-6 and liver stiffness also persisted (r = 0.340, p = 0.0342); however, the association between sgp130 and fat content disappeared (r = −0.003, p = 0.985) and a positive correlation between plasma sIL-6R and fat content (r = 0.358, p = 0.0223) was revealed. While this showed that general relationships between IL-6 signaling components and liver volume and stiffness in MASH were not significantly influenced by a history of HCC, this analysis promoted us to evaluate whether associations were dependent on other NAFLD comorbidities.

Stepwise forward regression analysis was performed to explore the influence of age, sex, BMI, and the other diseases associated with metabolic syndrome (diabetes, hyperlipidemia, hypertension, or previous HCC) on associations between IL-6 components and MASH severity (liver fat, volume and stiffness) (**Table 2**). Adjustment for any of these factors eliminated the associations of plasma IL-6 with all MRE/MRI measures of liver disease. On the other hand, plasma sIL-6R predicted liver volume independent of age, sex, BMI, and hyperlipidemia, hypertension or previous HCC history, but the association was influenced by diabetes status. Plasma sgp130 predicted liver stiffness independent of age, sex, BMI and any comorbidity measured, including diabetes **(Table 2)**.

**Table 2.**
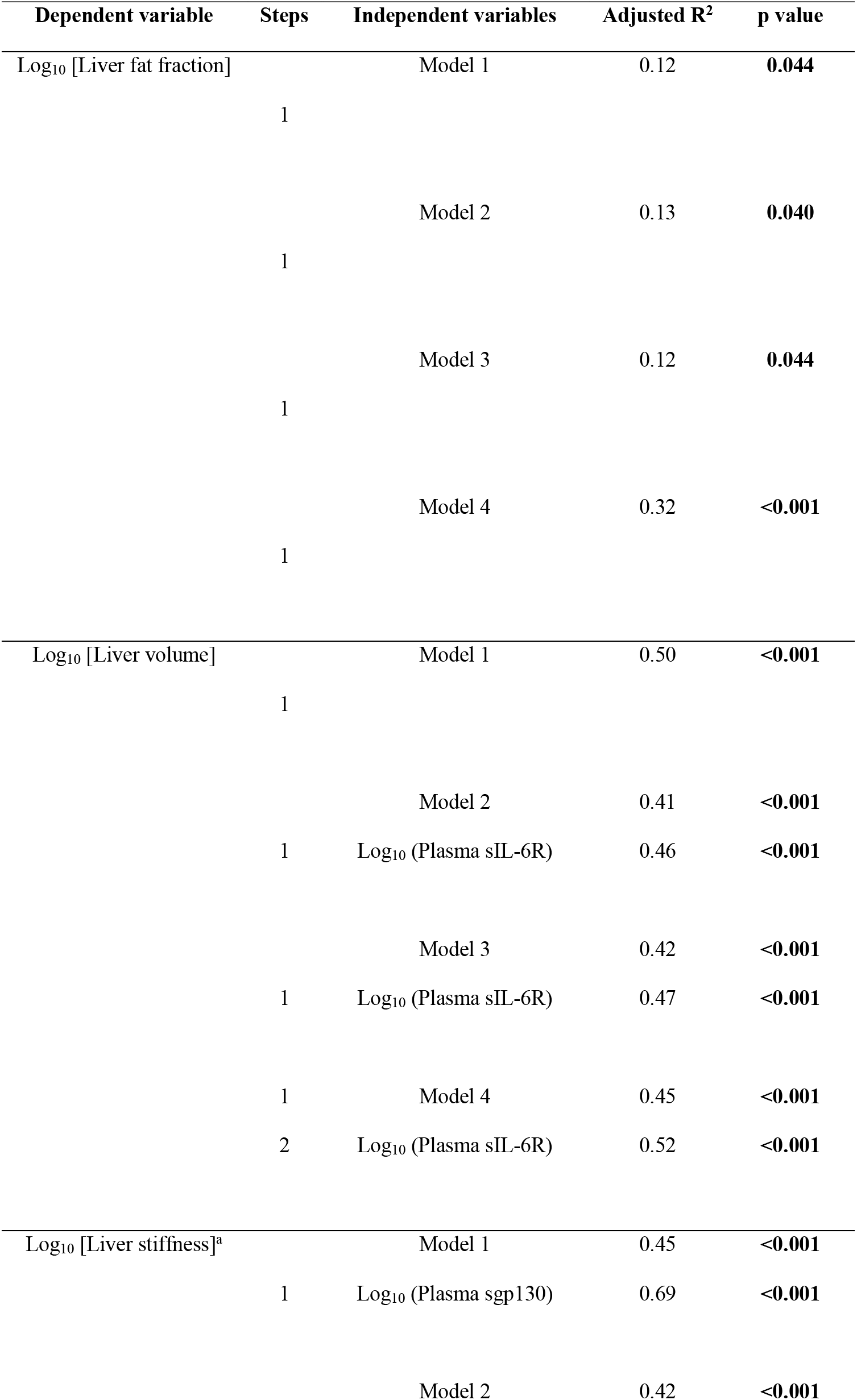
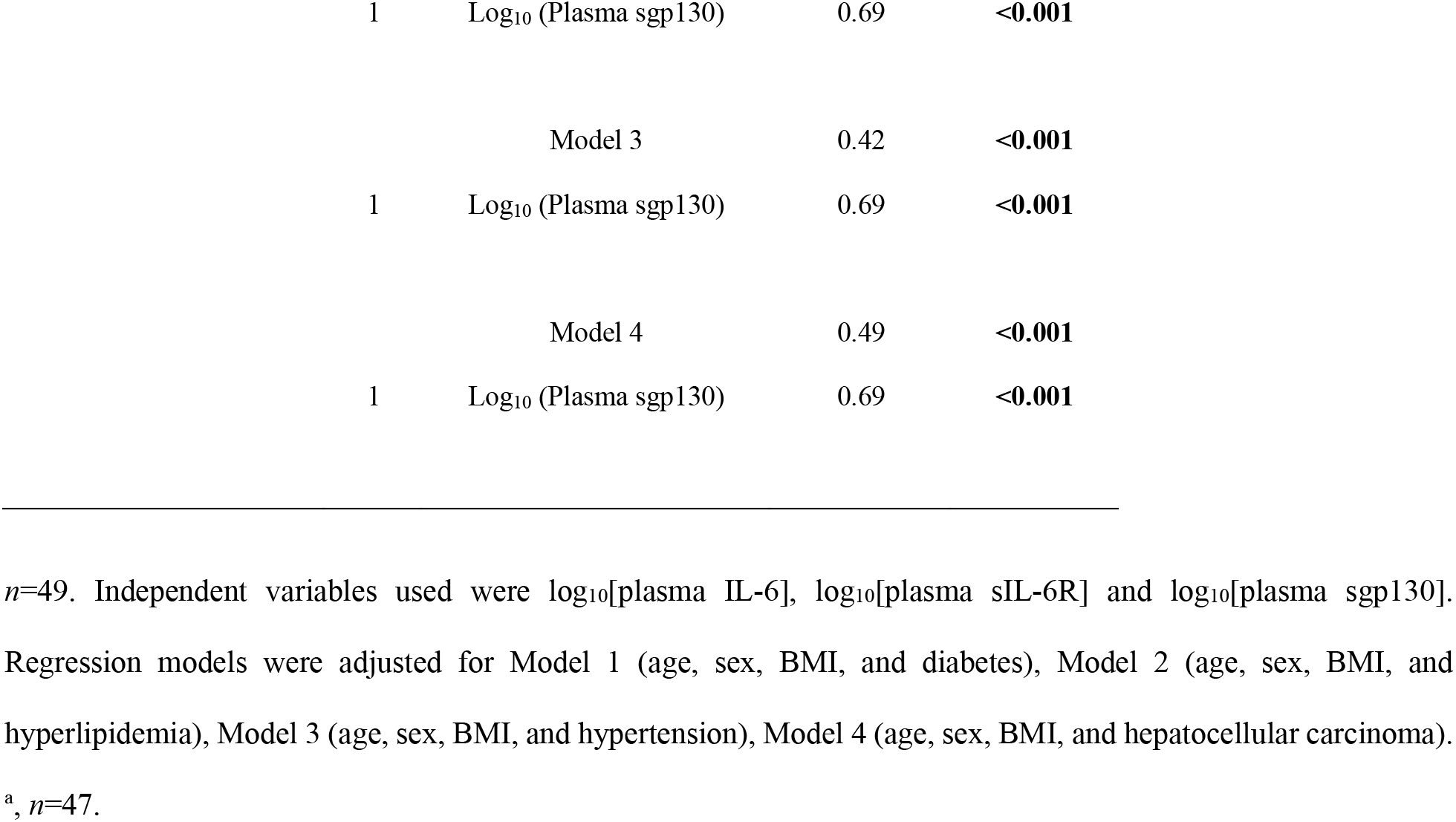
Prediction of liver fat fraction, liver volume and liver stiffness by plasma IL-6 related cytokines.

| Dependent variable | Steps | Independent variables | Adjusted R<sup>2</sup> | p value |
| --- | --- | --- | --- | --- |
| Log<sub>10</sub> [Liver fat fraction] |  | Model 1 | 0.12 | **0.044** |
|  | 1 |  |  |  |
|  |  | Model 2 | 0.13 | **0.040** |
|  | 1 |  |  |  |
|  |  | Model 3 | 0.12 | **0.044** |
|  | 1 |  |  |  |
|  |  | Model 4 | 0.32 | **<0.001** |
|  | 1 |  |  |  |
| Log<sub>10</sub> [Liver volume] |  | Model 1 | 0.50 | **<0.001** |
|  | 1 |  |  |  |
|  |  | Model 2 | 0.41 | **<0.001** |
|  | 1 | Log<sub>10</sub> (Plasma sIL-6R) | 0.46 | **<0.001** |
|  |  | Model 3 | 0.42 | **<0.001** |
|  | 1 | Log<sub>10</sub> (Plasma sIL-6R) | 0.47 | **<0.001** |
|  | 1 | Model 4 | 0.45 | **<0.001** |
|  | 2 | Log<sub>10</sub> (Plasma sIL-6R) | 0.52 | **<0.001** |
| Log<sub>10</sub> [Liver stiffness]<sup>a</sup> |  | Model 1 | 0.45 | **<0.001** |
|  | 1 | Log<sub>10</sub> (Plasma sgp130) | 0.69 | **<0.001** |
|  |  | Model 2 | 0.42 | **<0.001** |

|  |  |  |  |
| --- | --- | --- | --- |
| 1 | Log<sub>10</sub> (Plasma sgp130) | 0.69 | **<0.001** |
|  | Model 3 | 0.42 | **<0.001** |
| 1 | Log<sub>10</sub> (Plasma sgp130) | 0.69 | **<0.001** |
|  | Model 4 | 0.49 | **<0.001** |
| 1 | Log<sub>10</sub> (Plasma sgp130) | 0.69 | **<0.001** |

*n*=49. Independent variables used were log<sub>10</sub>[plasma IL-6], log<sub>10</sub>[plasma sIL-6R] and log<sub>10</sub>[plasma sgp130]. Regression models were adjusted for Model 1 (age, sex, BMI, and diabetes), Model 2 (age, sex, BMI, and hyperlipidemia), Model 3 (age, sex, BMI, and hypertension), Model 4 (age, sex, BMI, and hepatocellular carcinoma). <sup>a</sup>, *n*=47.

Histological evaluation of liver biopsy (steatosis, active liver inflammation and liver fibrosis) is the gold standard to diagnose MASH and define the range of disease severity. Histological scoring data (steatosis grade, activity score or fibrosis stage) were available from biopsies performed 6-12 months prior to imaging and blood collection. Plasma IL-6, sIL-6R or sgp130 were not significantly different between subjects with varying MASH severity by histological grading (**Supplementary Figure 3**). While this appears inconsistent with our findings above using imaging, there may have been changes in liver health during the time between liver biopsy and blood collection. Consistent with this possibility, there were also no correlations between histological scores and plasma ALT **(Supplementary Figure 4A, 4B, 4C)** or liver stiffness **(Supplementary Figure 4D, 4E, 4F)**.

In contrast, imaging and blood collection were performed on the same day, and multiple serum predictors of liver disease showed strong correlations with MRI/MRE measured liver stiffness, including INR-PT (r = 0.61, p < 0.0001), platelet count (r = −0.59, p < 0.0001), ALP (r = 0.55, p < 0.0001), GGT (r = 0.67, p < 0.0001), albumin (r = −0.52, p < 0.0001) and CK-18 (r = 0.38, p = 0.039) (**Table 3**). In addition, liver stiffness correlated strongly with all fibrosis risk scores. For these reasons, we focused on imaging-based measurements of liver disease severity for this cohort. We noted that the association of liver stiffness with plasma sgp130 was stronger than all other metabolic risk factors, liver disease scores or individual predictors measured (**Table 3**). These data suggest that the level of sgp130 in plasma is tightly associated with the extent of liver damage and stage of liver disease.

**Table 3.**
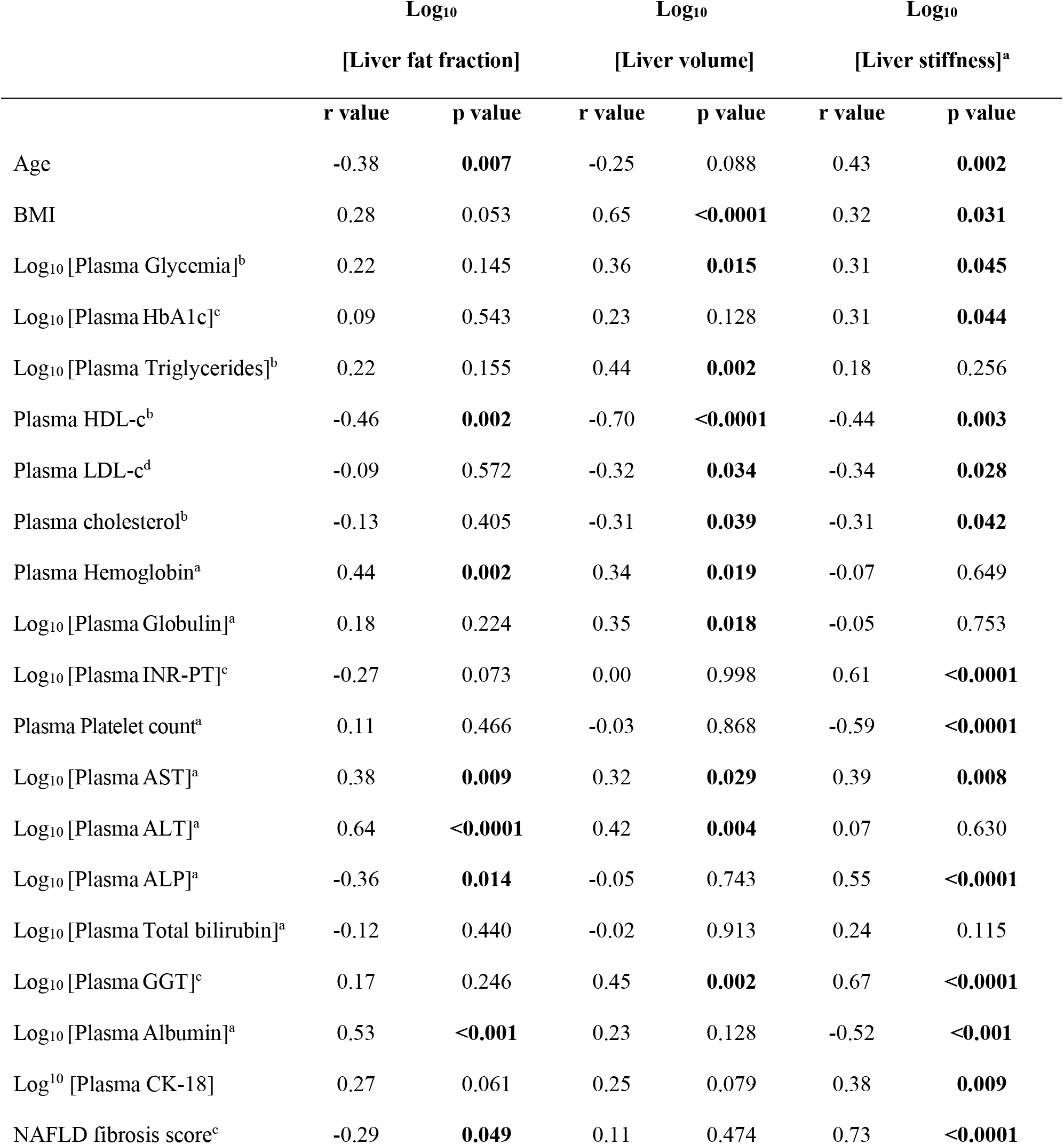
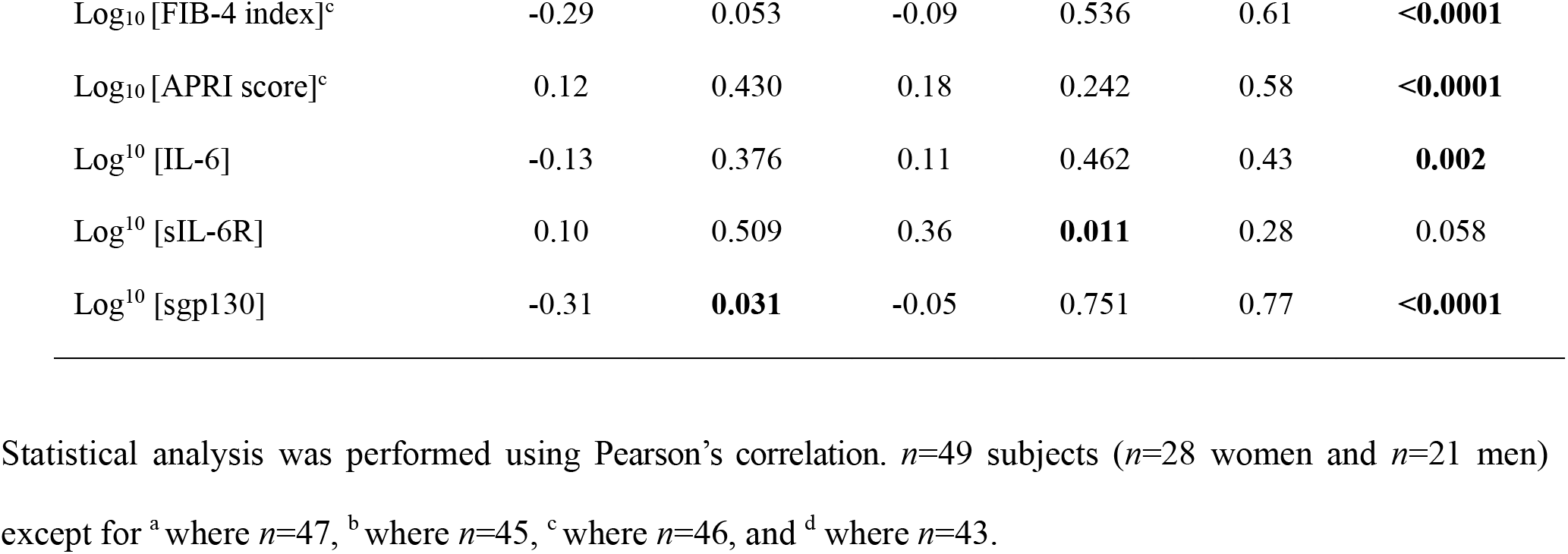
Correlations between common predictors of liver stiffness/fibrosis, liver risk scores and components of the IL-6 pathway with MRI/MRE measures of liver stiffness, liver fat fraction and liver volume in MASH subjects.

|  | Log<sub>10</sub><br>[Liver fat fraction] |  | Log<sub>10</sub><br>[Liver volume] |  | Log<sub>10</sub><br>[Liver stiffness]<sup>a</sup> |  |
| --- | --- | --- | --- | --- | --- | --- |
|  | r value | p value | r value | p value | r value | p value |
| Age | -0.38 | **0.007** | -0.25 | 0.088 | 0.43 | **0.002** |
| BMI | 0.28 | 0.053 | 0.65 | **<0.0001** | 0.32 | **0.031** |
| Log<sub>10</sub> [Plasma Glycemia]<sup>b</sup> | 0.22 | 0.145 | 0.36 | **0.015** | 0.31 | **0.045** |
| Log<sub>10</sub> [Plasma HbA1c]<sup>c</sup> | 0.09 | 0.543 | 0.23 | 0.128 | 0.31 | **0.044** |
| Log<sub>10</sub> [Plasma Triglycerides]<sup>b</sup> | 0.22 | 0.155 | 0.44 | **0.002** | 0.18 | 0.256 |
| Plasma HDL-c<sup>b</sup> | -0.46 | **0.002** | -0.70 | **<0.0001** | -0.44 | **0.003** |
| Plasma LDL-c<sup>d</sup> | -0.09 | 0.572 | -0.32 | **0.034** | -0.34 | **0.028** |
| Plasma cholesterol<sup>b</sup> | -0.13 | 0.405 | -0.31 | **0.039** | -0.31 | **0.042** |
| Plasma Hemoglobin<sup>a</sup> | 0.44 | **0.002** | 0.34 | **0.019** | -0.07 | 0.649 |
| Log<sub>10</sub> [Plasma Globulin]<sup>a</sup> | 0.18 | 0.224 | 0.35 | **0.018** | -0.05 | 0.753 |
| Log<sub>10</sub> [Plasma INR-PT]<sup>c</sup> | -0.27 | 0.073 | 0.00 | 0.998 | 0.61 | **<0.0001** |
| Plasma Platelet count<sup>a</sup> | 0.11 | 0.466 | -0.03 | 0.868 | -0.59 | **<0.0001** |
| Log<sub>10</sub> [Plasma AST]<sup>a</sup> | 0.38 | **0.009** | 0.32 | **0.029** | 0.39 | **0.008** |
| Log<sub>10</sub> [Plasma ALT]<sup>a</sup> | 0.64 | **<0.0001** | 0.42 | **0.004** | 0.07 | 0.630 |
| Log<sub>10</sub> [Plasma ALP]<sup>a</sup> | -0.36 | **0.014** | -0.05 | 0.743 | 0.55 | **<0.0001** |
| Log<sub>10</sub> [Plasma Total bilirubin]<sup>a</sup> | -0.12 | 0.440 | -0.02 | 0.913 | 0.24 | 0.115 |
| Log<sub>10</sub> [Plasma GGT]<sup>c</sup> | 0.17 | 0.246 | 0.45 | **0.002** | 0.67 | **<0.0001** |
| Log<sub>10</sub> [Plasma Albumin]<sup>a</sup> | 0.53 | **<0.001** | 0.23 | 0.128 | -0.52 | **<0.001** |
| Log<sub>10</sub> [Plasma CK-18] | 0.27 | 0.061 | 0.25 | 0.079 | 0.38 | **0.009** |
| NAFLD fibrosis score<sup>c</sup> | -0.29 | **0.049** | 0.11 | 0.474 | 0.73 | **<0.0001** |
| Log<sub>10</sub> [FIB-4 index]<sup>c</sup> | -0.29 | 0.053 | -0.09 | 0.536 | 0.61 | **<0.0001** |
| Log<sub>10</sub> [APRI score]<sup>c</sup> | 0.12 | 0.430 | 0.18 | 0.242 | 0.58 | **<0.0001** |
| Log<sup>10</sup> [IL-6] | -0.13 | 0.376 | 0.11 | 0.462 | 0.43 | **0.002** |
| Log<sup>10</sup> [sIL-6R] | 0.10 | 0.509 | 0.36 | **0.011** | 0.28 | 0.058 |
| Log<sup>10</sup> [sgp130] | -0.31 | **0.031** | -0.05 | 0.751 | 0.77 | **<0.0001** |

Statistical analysis was performed using Pearson's correlation. $n=49$ subjects ( $n=28$ women and $n=21$ men) except for <sup>a</sup> where $n=47$ , <sup>b</sup> where $n=45$ , <sup>c</sup> where $n=46$ , and <sup>d</sup> where $n=43$ .

### Plasma sgp130 predicts the severity of liver stiffness better than currently used plasma predictors of liver disease or calculated liver risk scores in patients with MASH

We next evaluated the strength of relationships between plasma components of the IL-6 pathway and: 1) metabolic risk factors (age, BMI, plasma glucose, glycated hemoglobin and lipids); 2) common risk scores used to estimate severity of the liver disease (NAFLD fibrosis score, FIB-4 index and APRI score); and, 3) individual plasma parameters used to estimate NASH risk/severity and calculate risk scores **(Supplementary Table 1)**. Plasma IL-6 correlated with common plasma predictors of liver disease including ALP (r = 0.47, p = 0.001), GGT (r = 0.30, p = 0.044), and albumin (r = −0.56, p < 0.0001), as well as scores used to assess MAFLD severity (NAFLD fibrosis score and FIB-4 index, r = 0.50, p = 0.001; r = 0.37, p = 0.011 respectively). In contrast, plasma sIL-6R correlated more readily with metabolic risk factors including glycemia (r = 0.55, p < 0.0001) and triglycerides (r = 0.37, p = 0.011) compared to other IL-6 components, while also correlating with the liver damage marker GGT (r = 0.50, p < 0.0001) and fibrosis risk scores. Plasma sgp130 correlated very strongly with multiple markers of liver damage including platelet count, INR-PT (r = 0.65, p < 0.0001), ALP (r = 0.62, p < 0.0001), GGT (r = 0.55, p < 0.0001), albumin (r = −0.66, p < 0.0001) and all fibrosis risk scores, but not with glycemia or lipid parameters. These data are in line with IL-6 signaling being a major player in metabolic disease (15, 17, 18, 26, 37–39) and suggest that different components of the IL-6 pathway (i.e. ligand and soluble receptors) may be differentially involved in the metabolic versus inflammatory aspects of fatty liver disease.

Given the strength of the associations of the three IL-6 signaling mediators with image-based measurements of liver disease and multiple markers of metabolic liver disease, we used stepwise linear regression analysis to identify the best predictors of liver fat, volume and stiffness, without or with adjustment for sex (**Table 4**). Sex was considered as a variable since many metabolic diseases including NAFLD and NASH are influenced by sex, and the liver is one of the most sexually dimorphic tissues (40–42). Independent parameters entered in the model were: the 3 components of the IL-6 pathway (IL-6, sIL-6R and sgp130); plasma parameters related to liver disease (globulin, INR-PT, ALP, total bilirubin and GGT); and liver disease scores (NAFLD fibrosis score, FIB-4 index and APRI score). Age and BMI were not adjusted for as they are used to calculate NAFLD fibrosis score and FIB-4 index.

**Table 4.**
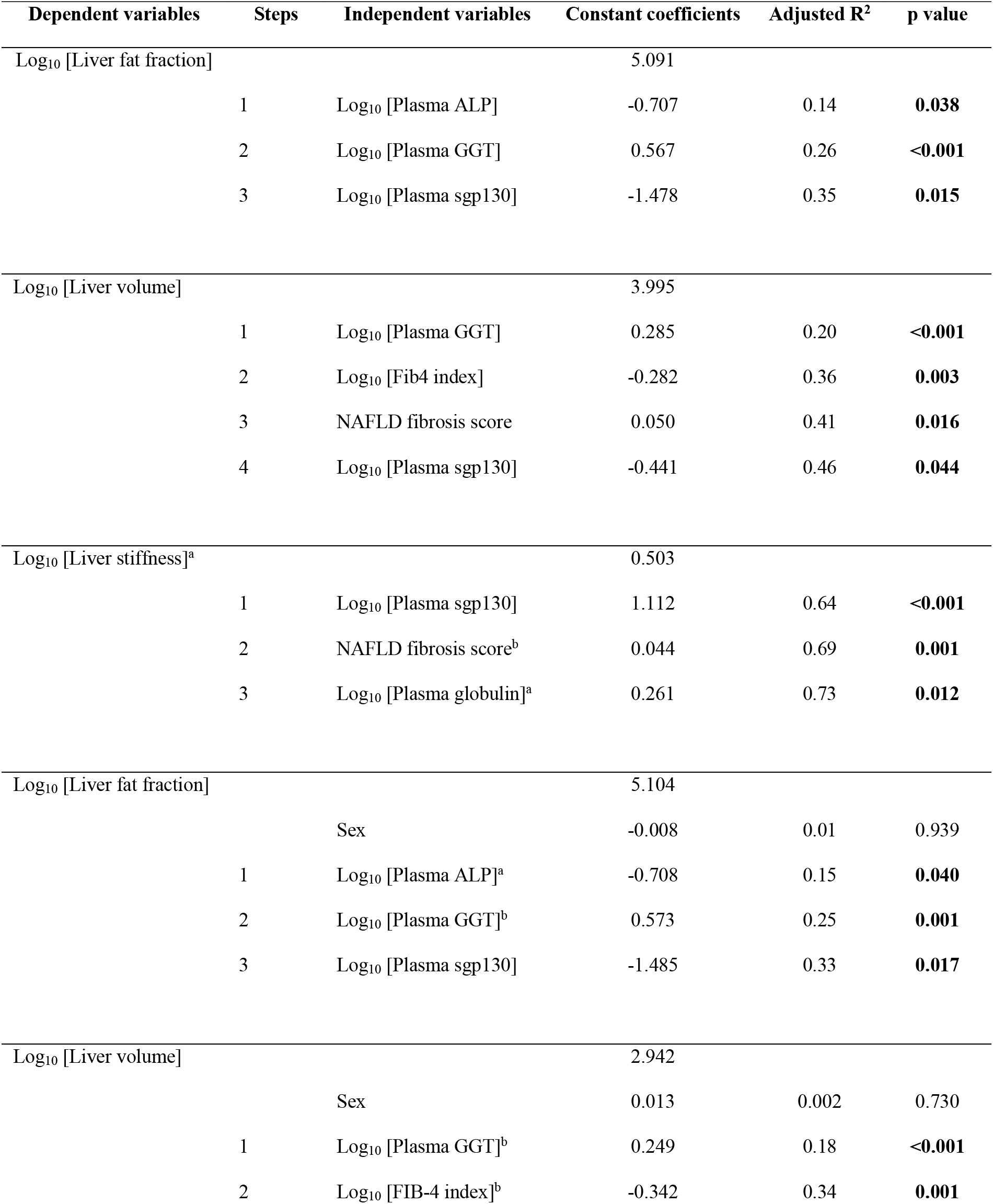
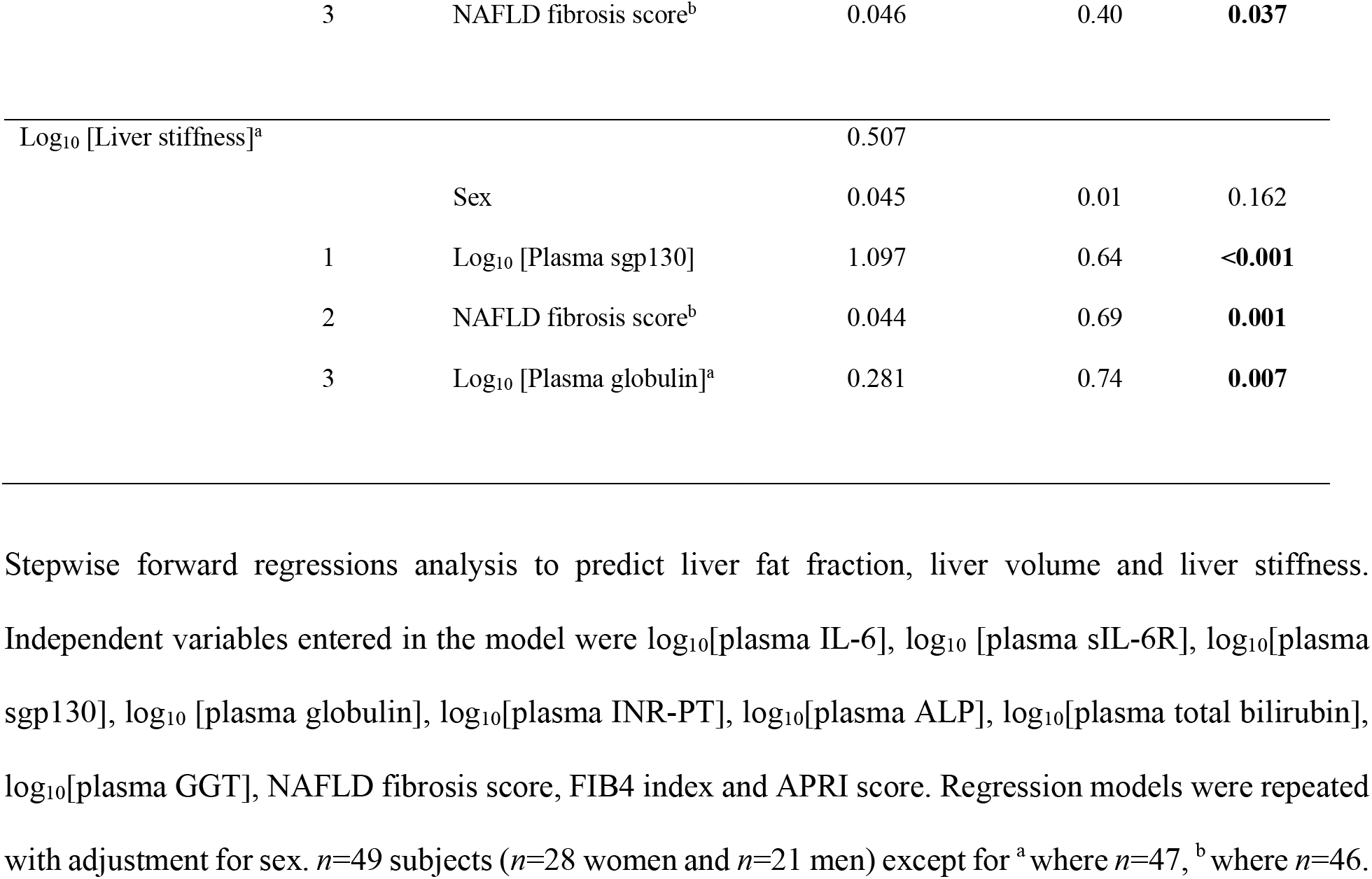
Stepwise linear regression to predict liver fat fraction, liver volume and liver stiffness in MASH patients.

Plasma ALP and GGT were the primary predictors of liver fat, predicting 26% of the intersubject variability, followed by plasma spg130, predicting an additional 9% (**Table 4**). All other parameters were excluded and adjustment for sex had little effect. Plasma sgp130 was retained in the regression model to predict liver volume after plasma GGT, FIB-4 index and NAFLD fibrosis score; however, adjustment for sex excluded sgp130 from this model. On the other hand, plasma sgp130 was the primary predictor of liver stiffness, alone explaining 64% of intersubject variability, while NAFLD fibrosis score and plasma globulin together explained an additional 9%. All other independent parameters were excluded and adjustment for sex had little effect to predict liver stiffness.

### Plasma sgp130 correlates with advanced liver disease in patients with morbid obesity

Anthropometric, metabolic, and clinical characteristics of patients with morbid obesity are presented in **Table 5**. In line with published correlations between obesity and NAFLD (1), 98% of these patients had hepatic steatosis. Among the population, 40.4% had diabetes, 81.9% had MAFLD, and 16.5% had MASH. However, despite extreme obesity (average BMI = 48), most were classified as MAFLD (not MASH) based on Bedossa scoring (43), with a high percentage of simple steatosis (70.6%, G0-G1), low activity score (81.9%, A0-A1), and low fibrosis stage (59.6%, F0-F1) assessed from the liver biopsies. Histological evaluation showed generally low scores for immune cell infiltration and hepatocyte ballooning, with average NAFLD activity scores (NAS) falling between 2-3 (**Table 5**). Taken together, the overall degree of liver disease severity was low in this cohort of morbid obesity, suggesting that this cohort had less active MASH compared to our first cohort at the time of liver biopsy.

**Table 5.**
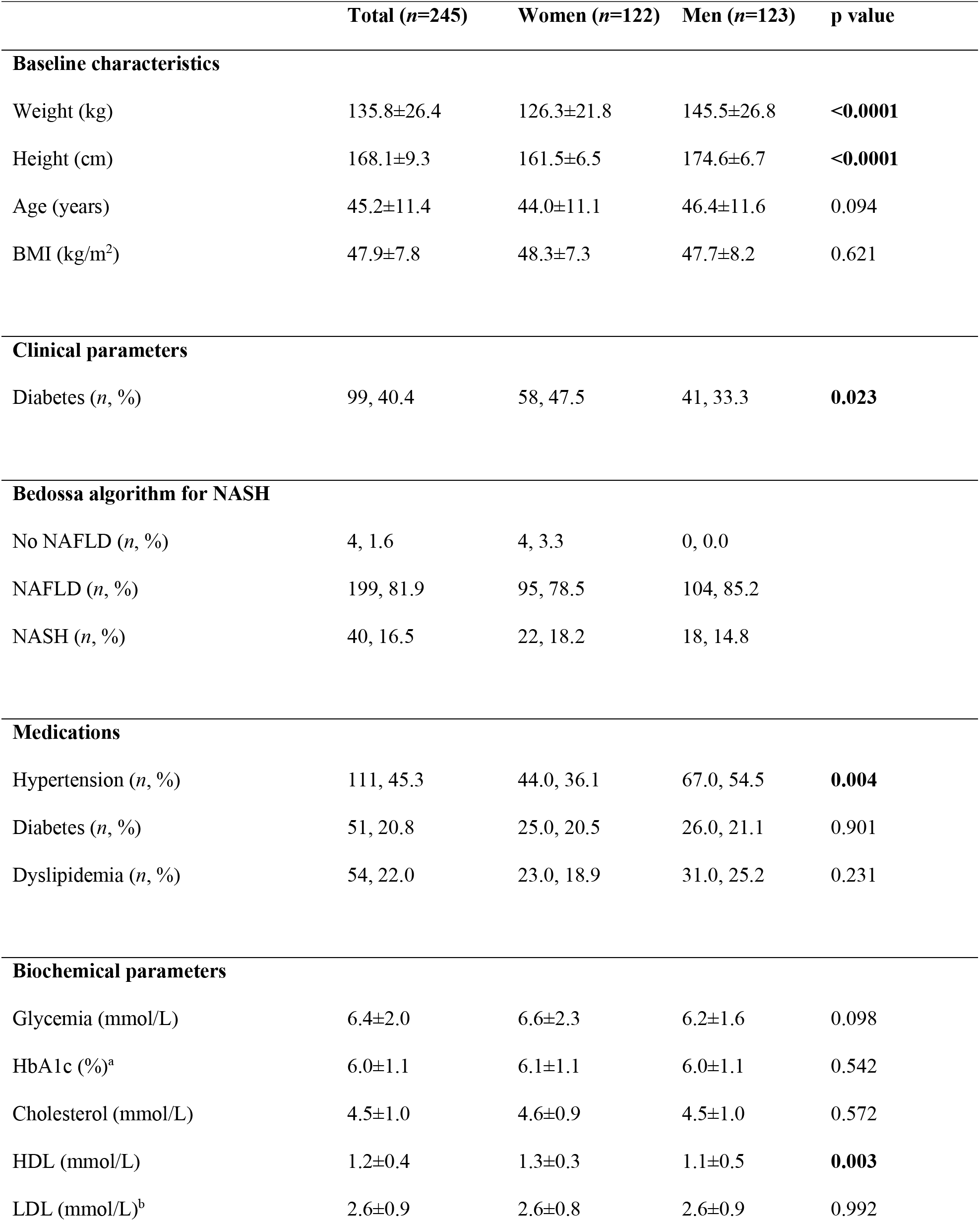
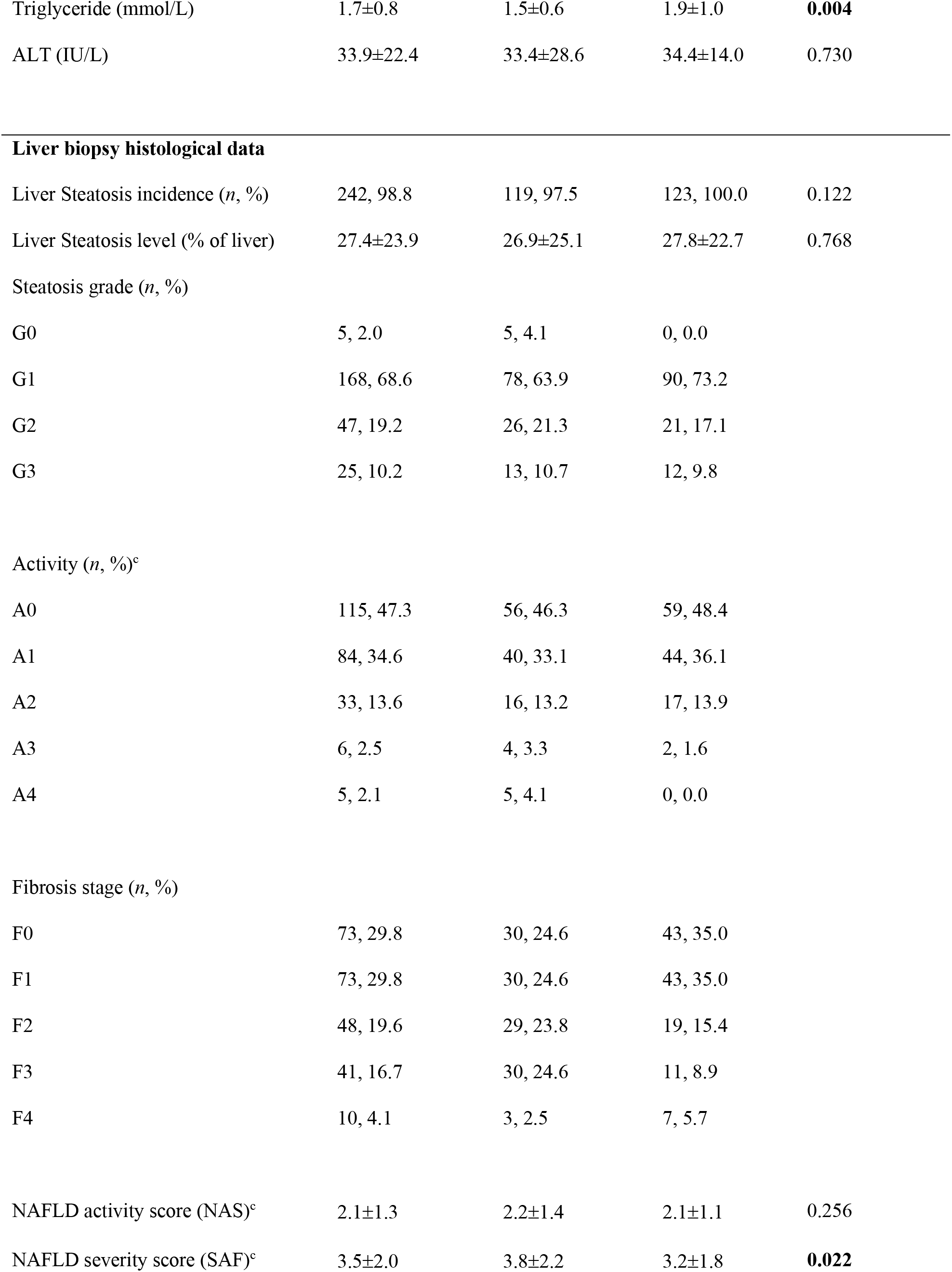
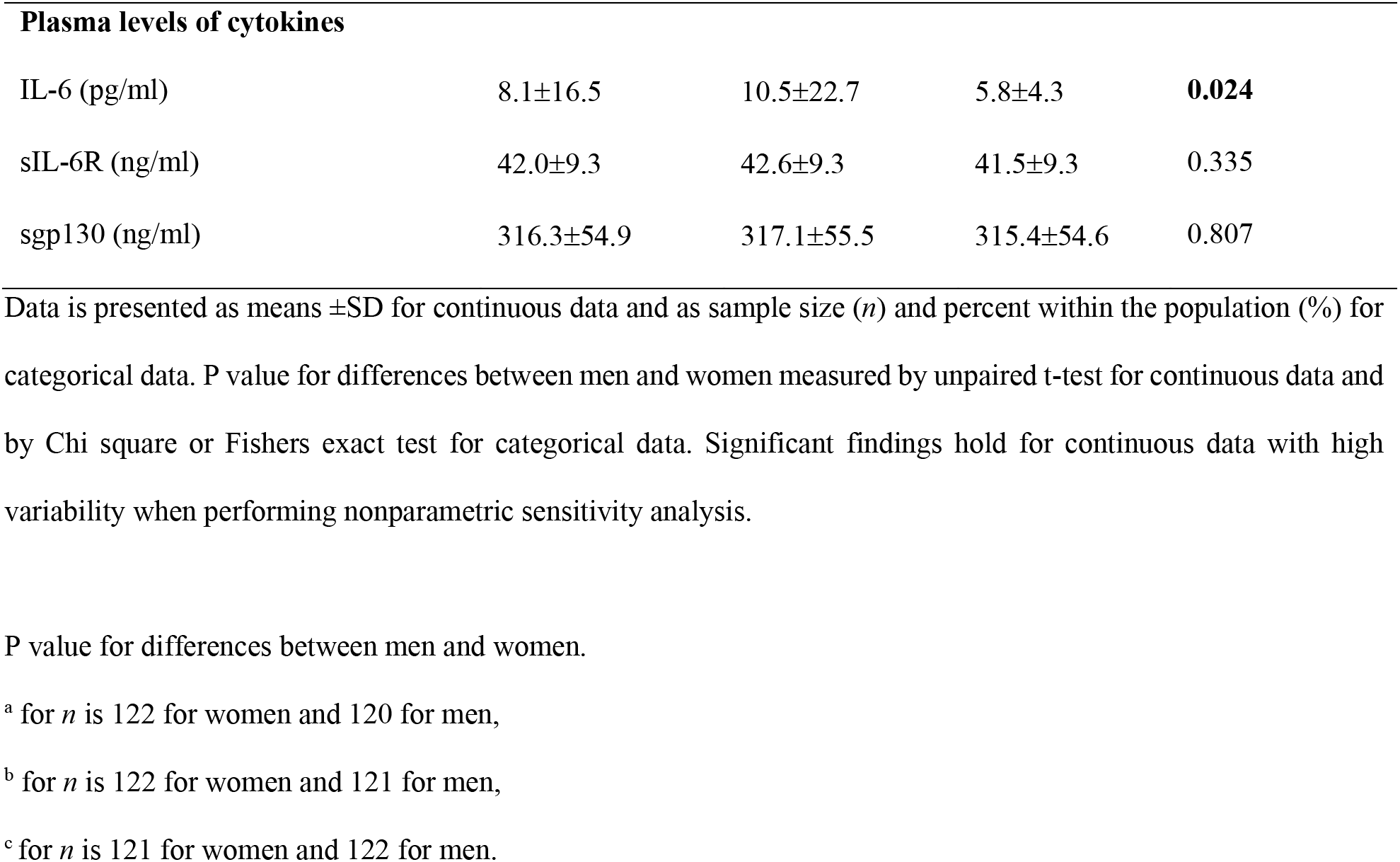
Anthropometric, metabolic and clinical characteristics of patients with morbid obesity.

|  | **Total (n=245)** | **Women (n=122)** | **Men (n=123)** | **p value** |
| --- | --- | --- | --- | --- |
| **Baseline characteristics** |  |  |  |  |
| Weight (kg) | 135.8±26.4 | 126.3±21.8 | 145.5±26.8 | **<0.0001** |
| Height (cm) | 168.1±9.3 | 161.5±6.5 | 174.6±6.7 | **<0.0001** |
| Age (years) | 45.2±11.4 | 44.0±11.1 | 46.4±11.6 | 0.094 |
| BMI (kg/m<sup>2</sup>) | 47.9±7.8 | 48.3±7.3 | 47.7±8.2 | 0.621 |
| **Clinical parameters** |  |  |  |  |
| Diabetes (n, %) | 99, 40.4 | 58, 47.5 | 41, 33.3 | **0.023** |
| **Bedossa algorithm for NASH** |  |  |  |  |
| No NAFLD (n, %) | 4, 1.6 | 4, 3.3 | 0, 0.0 |  |
| NAFLD (n, %) | 199, 81.9 | 95, 78.5 | 104, 85.2 |  |
| NASH (n, %) | 40, 16.5 | 22, 18.2 | 18, 14.8 |  |
| **Medications** |  |  |  |  |
| Hypertension (n, %) | 111, 45.3 | 44.0, 36.1 | 67.0, 54.5 | **0.004** |
| Diabetes (n, %) | 51, 20.8 | 25.0, 20.5 | 26.0, 21.1 | 0.901 |
| Dyslipidemia (n, %) | 54, 22.0 | 23.0, 18.9 | 31.0, 25.2 | 0.231 |
| **Biochemical parameters** |  |  |  |  |
| Glycemia (mmol/L) | 6.4±2.0 | 6.6±2.3 | 6.2±1.6 | 0.098 |
| HbA1c (%)<sup>a</sup> | 6.0±1.1 | 6.1±1.1 | 6.0±1.1 | 0.542 |
| Cholesterol (mmol/L) | 4.5±1.0 | 4.6±0.9 | 4.5±1.0 | 0.572 |
| HDL (mmol/L) | 1.2±0.4 | 1.3±0.3 | 1.1±0.5 | **0.003** |
| LDL (mmol/L)<sup>b</sup> | 2.6±0.9 | 2.6±0.8 | 2.6±0.9 | 0.992 |
| Triglyceride (mmol/L) | 1.7±0.8 | 1.5±0.6 | 1.9±1.0 | **0.004** |
| ALT (IU/L) | 33.9±22.4 | 33.4±28.6 | 34.4±14.0 | 0.730 |
| **Liver biopsy histological data** |  |  |  |  |
| Liver Steatosis incidence (*n*, %) | 242, 98.8 | 119, 97.5 | 123, 100.0 | 0.122 |
| Liver Steatosis level (% of liver) | 27.4±23.9 | 26.9±25.1 | 27.8±22.7 | 0.768 |

**Steatosis grade (*n*, %)**
|  |  |  |  |
| --- | --- | --- | --- |
| G0 | 5, 2.0 | 5, 4.1 | 0, 0.0 |
| G1 | 168, 68.6 | 78, 63.9 | 90, 73.2 |
| G2 | 47, 19.2 | 26, 21.3 | 21, 17.1 |
| G3 | 25, 10.2 | 13, 10.7 | 12, 9.8 |

**Activity (*n*, %)<sup>c</sup>**
|  |  |  |  |
| --- | --- | --- | --- |
| A0 | 115, 47.3 | 56, 46.3 | 59, 48.4 |
| A1 | 84, 34.6 | 40, 33.1 | 44, 36.1 |
| A2 | 33, 13.6 | 16, 13.2 | 17, 13.9 |
| A3 | 6, 2.5 | 4, 3.3 | 2, 1.6 |
| A4 | 5, 2.1 | 5, 4.1 | 0, 0.0 |

**Fibrosis stage (*n*, %)**
|  |  |  |  |
| --- | --- | --- | --- |
| F0 | 73, 29.8 | 30, 24.6 | 43, 35.0 |
| F1 | 73, 29.8 | 30, 24.6 | 43, 35.0 |
| F2 | 48, 19.6 | 29, 23.8 | 19, 15.4 |
| F3 | 41, 16.7 | 30, 24.6 | 11, 8.9 |
| F4 | 10, 4.1 | 3, 2.5 | 7, 5.7 |
| NAFLD activity score (NAS)<sup>c</sup> | 2.1±1.3 | 2.2±1.4 | 2.1±1.1 | 0.256 |
| NAFLD severity score (SAF)<sup>c</sup> | 3.5±2.0 | 3.8±2.2 | 3.2±1.8 | **0.022** |

| **Plasma levels of cytokines** |  |  |  |  |
| --- | --- | --- | --- | --- |
| IL-6 (pg/ml) | 8.1±16.5 | 10.5±22.7 | 5.8±4.3 | **0.024** |
| sIL-6R (ng/ml) | 42.0±9.3 | 42.6±9.3 | 41.5±9.3 | 0.335 |
| sgp130 (ng/ml) | 316.3±54.9 | 317.1±55.5 | 315.4±54.6 | 0.807 |

Data is presented as means ±SD for continuous data and as sample size (*n*) and percent within the population (%) for categorical data. P value for differences between men and women measured by unpaired t-test for continuous data and by Chi square or Fishers exact test for categorical data. Significant findings hold for continuous data with high variability when performing nonparametric sensitivity analysis.
P value for differences between men and women.
<sup>a</sup> for *n* is 122 for women and 120 for men,
<sup>b</sup> for *n* is 122 for women and 121 for men,
<sup>c</sup> for *n* is 121 for women and 122 for men.

Average plasma IL-6, sIL-6R and spg130 were all above levels reported in healthy subjects (30, 31) **(Table 5)**. Similar to the MASH cohort, plasma IL-6 (p = 0.029), sIL-6R (p = 0.043) and sgp130 (p < 0.0001) were higher in subjects with diabetes **(Supplementary Figure 5A-C)**. Consistent with being a risk factor for advanced disease, subjects with diabetes had higher steatosis grade, activity score and fibrosis stage determined using liver biopsy samples **(Supplementary Figure 6A-C)**; however, BMI did not vary with any of these histological parameters **(Supplementary Figure 6D-F)**. After adjusting for age, sex, BMI and diabetes, neither IL-6, sIL-6R nor sgp130 corresponded with steatosis grade or activity score. However, plasma sgp130 was higher in subjects with an F4 fibrosis stage compared to subjects at all other stages (F0-F3) (p = 0.0001) **(Figure 3)**. In contrast, higher plasma ALT corresponded with G3 steatosis grade (p = 0.019), A1 activity score (p = 0.0249), and F2 fibrosis (p = 0.0149), but levels plateaued quickly at early stages **(Supplementary Figure 6G, H, I)**. Other serum parameters of liver damage were not available from this biobank. Thus, in this cohort, our analysis suggests that increased circulating sgp130 corresponded to and was specific for advanced liver fibrosis.

**Figure 3.**
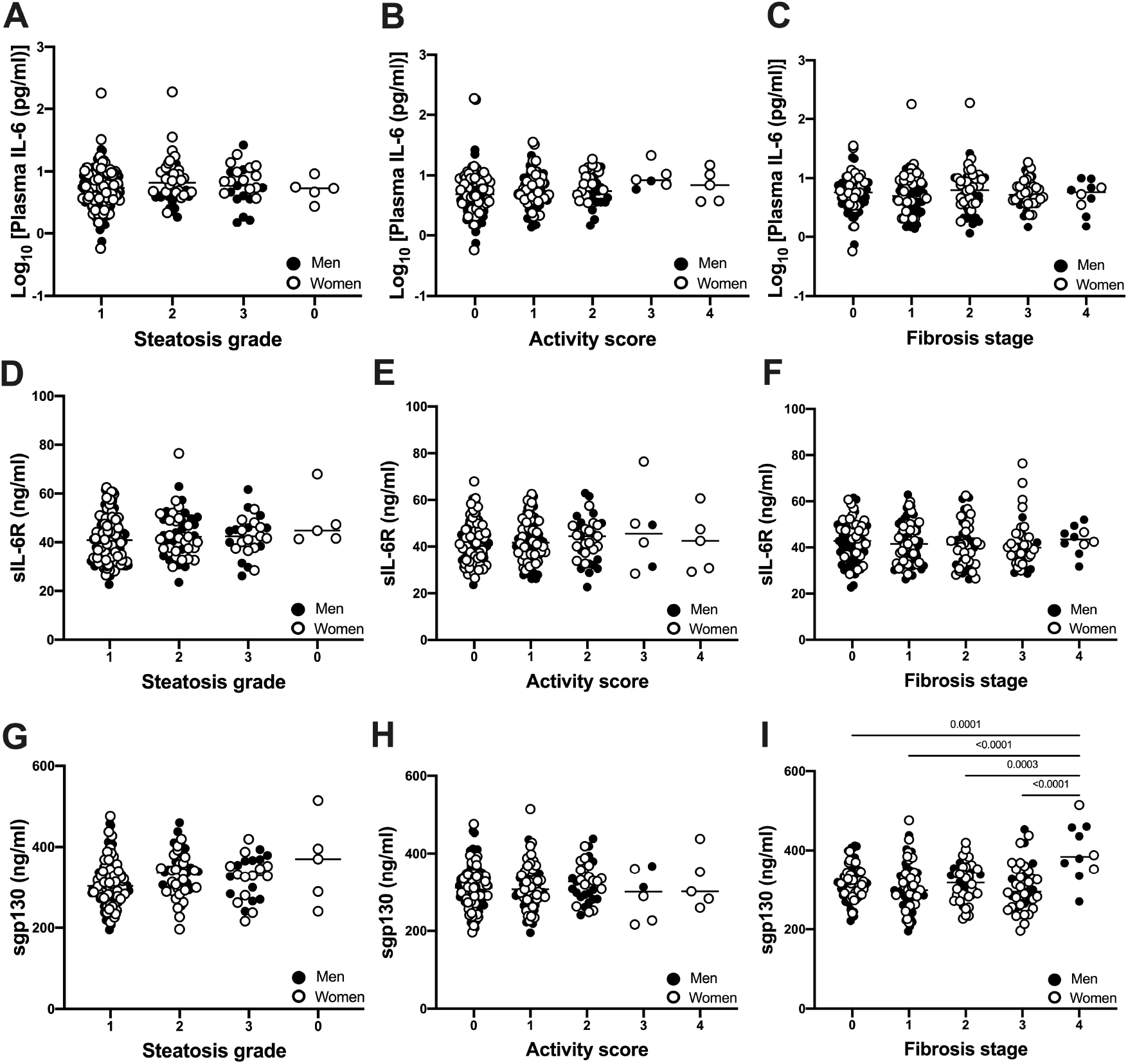
Plasma concentrations of IL-6 (A, B, C), sIL-6R (D, E, F) and sgp130 (G, H, I) in patients with morbid obesity compared among liver biopsy measures of steatosis grade, activity score and fibrosis stage. Data presented for subjects with steatosis grade G0 (*n*=5), G1 (*n*=168), G2 (*n*=47), F3 (*n*=41), F4 (*n*=10), subjects with activity score A0 (*n*=115), A1 (*n*=84), A2 (*n*=33), A3 (*n*=6), A4 (*n*=5) and subjects with fibrosis stage F0 (*n*=73), F1 (*n*=73), F2 (*n*=48), F3 (*n*=41), F4 (*n*=25). Analysis was conducted using 1-way ANOVA with multiple comparisons adjusted for age, sex, BMI and diabetes.

## DISCUSSION

In this study we explored relationships between three components of the IL-6 signaling pathway (IL-6, sIL-6R and sgp130) and metabolic fatty liver disease evaluated by liver biopsy, MRE/MRI, plasma predictors and/or liver risk scores in two separate cohorts with diagnosed MASH or morbid obesity. We provide novel data showing that: 1) plasma concentrations of IL-6 transsignaling mediators are differentially increased in obesity, diabetes, hypertension and/or previous history of HCC in patients with MASH; 2) plasma sgp130 strongly predicts MRE/MRI measured liver stiffness in MASH, independent of age, sex, BMI and any comorbidity (diabetes, hypertension, hyperlipidemia or history of HCC), performing superior to many other plasma predictors and liver risk scores; 3) plasma sgp130 is increased in advanced fibrosis independent of age, sex, BMI and diabetes in patients with morbid obesity; and finally, 4) plasma sIL-6R predicts MRE/MRI measures of liver volume, independent of age, sex and BMI.

Circulating inflammatory factors that associate tightly with MAFLD are not well known. We found higher levels of circulating IL-6 and sgp130 in patients with MAFLD/MASH, and that all three circulating IL-6 transignaling components are further increased by concomitant diabetes. This is in line with previous associations of these factors with metabolic syndrome (17–20, 39). However, our new data reveal a strong link between plasma levels of these factors and fatty liver disease, a pathology often occurring concurrently with diabetes and other metabolic diseases. Our data support liver as a possible major source for circulating components of IL-6 transsignaling, and that transignaling may contribute to the development or progression of MAFLD and other metabolic diseases.

The role of IL-6 transsignaling on liver function is controversial. Activation of IL-6 transsignaling promotes tumor formation in a mouse model of HCC (44) and selective blockade of IL-6 transsignaling by sgp130 decreases liver regeneration following partial hepatectomy (45). These studies suggest that IL-6 transsignaling may promote hepatocyte proliferation. Ablation of total hepatic IL-6 signaling in mice causes steatosis and fibrosis (46), but the contribution of classical versus trans-IL-signaling is not clear. In line with their human data (46), we also show no significant association between plasma sIL-6R and liver fibrosis stages in our cohorts; however, we find circulating sIL-6R significantly associated with multiple metabolic aspects of MAFLD including glycemia and lipidemia, hepatic fat fraction, as well as total liver volume. Interestingly, larger liver volume was recently associated with a 3-fold increase in risk of all-cause mortality in NAFLD (47). While we observe strong associations of sgp130 with liver stiffness and advanced fibrosis in humans with NASH, sgp130 treatment does not impact NASH development in mice fed a western diet (48). More work is needed to determine whether local or systemic secretion of IL-6-transsignaling factors has a direct effect on the progression of MAFLD/MASH.

Using stepwise linear regression analysis, we show for the first time that plasma sgp130 alone is a strong predictor of liver stiffness in MASH with a wide range of disease severity, better than commonly used serum predictors and risk scores. When combined with NAFLD fibrosis score and plasma globulin, the predicative power of the model increased. Thus, we propose that plasma sgp130 (alone or in combination with NAFLD fibrosis score) could be an effective, non-invasive method to predict the severity of liver disease in patients with MASH. Identification of non-invasive, reliable predictors of hepatic inflammation and fibrosis, especially at early stages (F1 and F2), will facilitate diagnosis and possibly increase success rates of emerging interventions and treatment strategies. We present this as a hypothesis-generating study and recognize that a larger sample size is required to confirm IL-6 transsignaling proteins as biomarkers of MAFLD.

One limitation of our study is the use of different methods to quantify liver disease severity across cohorts. For the first, we relied mainly on MRI and MRE analysis to assess severity of liver disease, while for the second cohort we used scoring data from liver biopsies. Liver biopsy data was available for the MASH cohort; however, the sample was taken up to 12 months prior to the blood draw and disease severity may have changed over that time. Liver biopsy remains gold standard, yet accumulating studies show that MRI/MRE also has high diagnostic accuracy for liver fibrosis, that is similar to biopsy in subjects with NAFLD (33–35). Regardless, the two methods of assessing liver disease severity limited our ability to directly compare liver disease parameters (steatosis grade, activity scores, and fibrosis stage) between the cohorts.

Liver fibrosis and liver stiffness are also not interchangeable. Liver fibrosis in biopsies is scored based on collagen staining, while MRE-determined liver stiffness is influenced by influenced to varying degress by fibrosis, inflammation, and steatosis (33–35). This may explain discrepancies between sgp130 sensitivity to predict stiffness versus fibrosis across the two cohorts. Interestingly, if we consider this differences, our data could also suggest that spg130 is a sensitive marker of *active* liver inflammation and damage, but not a predictor of collagen deposition per se. Sgp130 is a component of an inflammatory signaling pathway, which could explain stronger correlation with liver stiffness versus collagen staining (a late consequence of damaging stimuli). In line with this theory, sgp130 was significantly increased only in late fibrosis stages determined by histology, while the strong linear correlation between plasma sgp130 and liver stiffness measured by MRI/MRE spanned across all stages of disease severity.

In conclusion, our data support that circulating components of the IL-6 signaling system may play a role in MAFLD/NASH pathogenesis and have potential to serve as sensitive predictors of liver damage associated with metabolic disease. Importantly, relationships between plasma sIL-6R and particularly spg130 with metabolic liver disease severity are independent of sex, age and BMI. Their individual associations with either metabolic or inflammatory aspects of MAFLD suggest interesting mechanistic roles in disease progression. Their strong associations with liver fat, volume and stiffness may also be useful for non-invasive monitoring of the early MAFLD to MASH transition.

## METHODS

### Patients with MASH

MASH had been previously confirmed by liver biopsies performed 6-12 months prior to MRI/MRE, according to the clinical standard of care using 16-G or 18-G core needles. Hematoxylin and eosin (H&E) slides were scored by liver pathologists, with fibrosis stage, inflammation grade, and steatosis grade assessed according to the NASH Clinical Research Network (NASH CRN) histological scoring system(49).

Subjects were aged 18 years and older, diagnosed with MASH or with HCC on a MASH background, able to undergo magnetic resonance imaging (MRI) without administration of a contrast agent and to understand instructions in either French or English. Subjects were excluded if they: had high alcohol consumption (>10 drinks/week for women and >15 for men); had liver disease other than MASH; were taking medications associated with steatosis (e.g. amiodarone, valproate, tamoxifen, methotrexate or corticosteroids); physically unable to fit in the MRI machine; had contraindications to MRI; or were pregnant or wished to be pregnant during the study-year.

Of the 493 patients, 133 patients were eligible for this study, of whom 89 were diagnosed with MASH and 44 with MASH and a previous history of HCC. Between May 2018 and June 2019, which represents six to twelve months after the liver biopsies and diagnosis of MASH, the 133 patients were invited back for an MRI/MRE scan to assess liver fat fraction, volume and liver stiffness. Among these, 49 subjects with MASH alone and 34 with HCC and MASH did not participate in the study for the following reasons: refusal (*n*=40), unreachable (*n*=25), cancellation (*n*=8), distance to the hospital (*n*=8), language issues (*n*=1) or other reasons (comorbidities including amputations) (*n*=1). Thus, this analysis includes 40 patients with NASH (28 women, 22 men) and 10 patients with MASH and a history of HCC (4 women and 6 men) from the original registry of 493 patients. All patients (*n*=50) signed a consent form to be included in this study, approved by the Centre hospitalier de l’Université de Montréal human ethics committee (IRB #17.031). A flowchart of inclusion/exclusion criteria used is shown in **Supplementary Figure 1**.

MRI/MRE examinations were performed using a 3.0 T clinical scanner (Skyra; Siemens Healthineers, Mountain View, California). Proton density fat fraction (PDFF), liver volume (voxels, cm^3^), and liver stiffness (Pa) were measured as quantitative predictors of liver fat, volume and fibrosis, respectively. Average PDFF values for the entire liver volume were obtained using the LiverLab package (Magnetom Aera, Software version VE11C, Siemens Healthcare GmbH, Erlangen, Germany). Liver stiffness measurements by MRE were performed according to previously described methods (50). Fasting plasma samples were collected on the day of the MRI/MRE and stored at −80°C until measurement of plasma IL-6 parameters and other predictors for calculation of the liver disease scores.

### Calculation of fasting NAFLD fibrosis score, FIB-4 index and APRI score

The following equations were used to calculate liver scores in the NASH population:

NAFLD fibrosis score = −1.675 + [0.037 x age (years)] + [0.094 x BMI (kg/m^2^)] + [1.13 × impaired fasting glucose/diabetes (yes=1, no=0)] + [0.99 × AST/ALT ratio] - [0.013 × platelet count (× 10^9^/L)] – [0.66 × albumin (g/dl)] (51).
Fibrosis-4 (FIB-4) index = [Age (years)] × [AST (U/L)] / [platelet count (10^9^/L) × sqrt (ALT) (U/L)] (52).
AST to platelet count ratio index (APRI): [AST (IU/L)/40 IU/L] / [platelet count (× 10^9^/L)]] × 100 (53).

### Patients with morbid obesity

Adult men and women (18 years and older) were selected from a registry of 4,781 patients undergoing bariatric surgery. Exclusion criteria were: having high alcohol consumption (>10 drinks/week for women and >15 for men); or having liver disease other than NASH (e.g. autoimmune hepatitis, Wilson disease, hemochromatosis or HBV, HCV or human immunodeficiency viruses). Thus, this retrospective analysis included a subpopulation of 245 subjects (123 men, 122 women) selected based on availability of serum and histological scoring data.

Random blood samples were collected on the night before bariatric surgery and stored immediately at −80°C until time of analysis. Sampling procedure and position were standardized among all surgeons. Liver samples were obtained by incisional biopsy of the left lobe and were not cauterized. Grading and staging of histological liver sections were performed using the protocol of Brunt et al (54) by pathologists blinded to the study objectives. Bedossa algorithm (43) was used to diagnose NASH, using liver biopsy histological scores for hepatocellular ballooning stage (0-2), lobular inflammation (0-2), steatosis grade (G0-G3), activity score (A0-A4) and fibrosis stage (F0-F4). These were also used to calculate NAFLD activity (NAS) and steatosis, activity and fibrosis (SAF) scores (43).

### Measurements of plasma IL-6, sIL-6R, sgp130, Cytokeratin-18 and Alanine aminotransferase (ALT)

Commercial ELISA kits were used to measure plasma concentrations of IL-6, sIL-6R and sgp130 (R&D systems, Human Quantikine ELISA kits, D6050, DR600, and DGP00 respectively) and Cytokeratin-18 (Peviva, M30 Apoptosense ELISA, 10011). For sgp130 and sIL-6R, samples were diluted 1:100, while for IL-6 and Cytokeratin-18, undiluted samples were used. ALT was measured in patients with morbid obesity using a commercial kit (SGPT liquid ALT reagent set, Pointe scientific A7526). All assays performed according to manufacturer’s instructions and quality controls/standard provided with kits were included. Sample analysis was blinded using subject identification code.

### Statistics

Data in Table 1 and Table 5 are presented as mean±SD for continuous variables and as the number of subjects (*n*) and percentage (%) within the subpopulation for categorial variables. Normality was evaluated using a Kolmogrov-Smirnov test. When normality failed, data was log transformed (log_10_). Outlier and influencer points were identified using SPSS. One subject in the NASH cohort was a strong influencer for plasma IL-6 and sIL-6R in all analysis. Thus, this subject was excluded from analyses. Unpaired t-test, one-way ANOVA and Pearson correlation were used to analyze parametric or log_10_ transformed data. Given the variability of some continuous data in tables 1 and 5, sensitivity analysis was performed using nonparametric Mann-Whitney *U* test for intergroup differences to validate significant findings. For categorical variables, chi-square test was used for count >5 in each cell, otherwise Fisher’s exact test was used.

Clinical endpoints used were liver fat fraction, volume and stiffness for the NASH cohort, and liver steatosis grade, activity score and fibrosis stage for the morbid obesity cohort. To assess relationships between circulating IL-6, sIL-6R and sgp130 and these endpoints, stepwise forward regression analysis was used to predict measurements of liver disease severity using log_10_ [plasma IL-6], log_10_ [plasma sIL-6R] and log_10_ [plasma sgp130] as independent variables with adjustment for age, sex, and BMI and comorbidities (diabetes, hyperlipidemia, hypertension or previous HCC history) for the NASH cohort. For the morbid obesity cohort, log_10_ [plasma IL-6], sIL-6R and sgp130 were used as independent variables with adjustment for age, sex, BMI and diabetes. For categorical variables (NASH cohort: liver steatosis stage, activity score and fibrosis stage. Morbid obesity cohort: liver steatosis stage, activity score and fibrosis stage, Bedossa NASH diagnosis, NAS and SAF score), univariate analysis was used with adjustment for age, sex, BMI and diabetes. Data was analyzed using IBM SPSS (Version 27) and GraphPad Prism (Version 8) and significance was set at *P* < 0.05.

### Study approval

#### Patients with MASH

Participants were selected from a registry of 493 patients with available liver biopsies collected under ethical approval (IRB: #15.147) at Centre hospitalier de l’Université de Montréal that was initiated in February 2016. All selected participants (n = 50) in the current study (IRB # 17.031) provided written, informed consent allowing preservation and subsequent use of their data. All participants provided written, informed consent allowing preservation and subsequent use of their plasma samples and data.

#### Patients with morbid obesity

Plasma samples and matching liver biopsies were obtained from the Biobank of the *Institut universitaire de cardiologie et de pneumologie de Québec – Université Laval* in compliance with Institutional Review Board-approved management policies initiated in 2002 and still ongoing. Liver biopsy samples were collected at the time of bariatric surgery under ethical approval (IRB: #1142).

## Data Availability

All data produced in the present study are available upon reasonable request to the authors

## Author Contributions

JLE and AT designed studies. CH and AB managed the NASH clinical study. L Biertho and AL designed and carried out the bariatric surgery study. AT, L Bilodeau, AG and C Baldwin acquired, analyzed and interpreted data. MF assisted with data analysis. JLE, L Bilodeau, AT, MF, ML, EG, MB, C Bémeur contributed intellectual content. CYC scored liver histological samples. AG, MF and JLE wrote the manuscript. All authors reviewed the manuscript.

## Acknowledgements

Authors acknowledge the invaluable collaboration of the surgery team, bariatric surgeons, and biobank staff of the IUCPQ. We would also like to thank Paule Bodson Clermont, Hannah Zhang, Mélissa Léveillé and Stewart Jeromson for their assistance.

## SUPPLEMENTARY MATERIAL

**Supplementary Figure 1.**
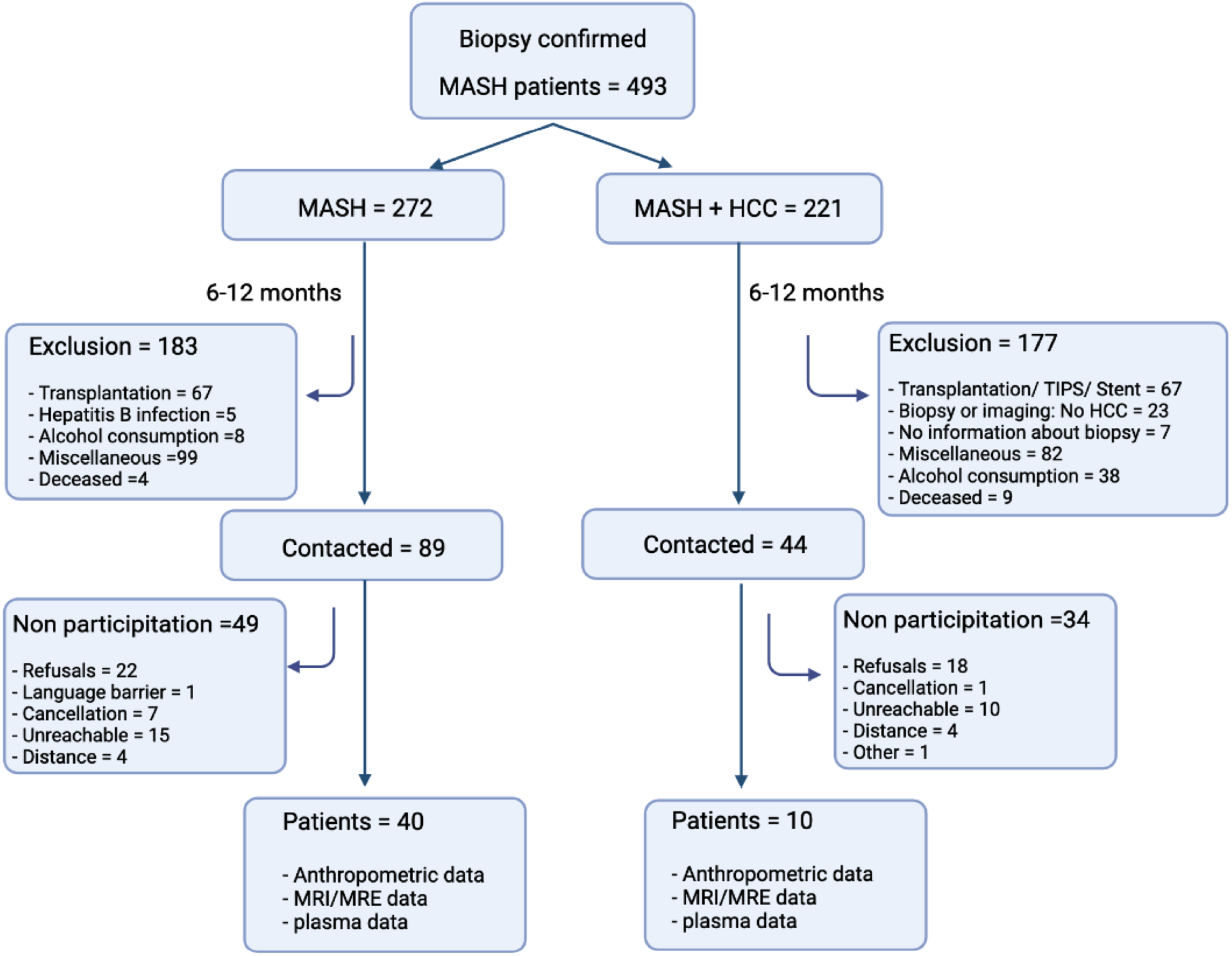
Study design for MASH cohort. Between May 2018 and June 2019, eligible patients with a history of biopsy-confirmed MASH were contacted to participate in our study. Flow chart details the inclusion and exclusion criteria used, as well as reasons for non-participation. Consenting patients (N=50) returned to the Centre hospitalier de l’Université de Montréal for MRE/MRI scan and blood draw.

**Supplementary Figure 2.**
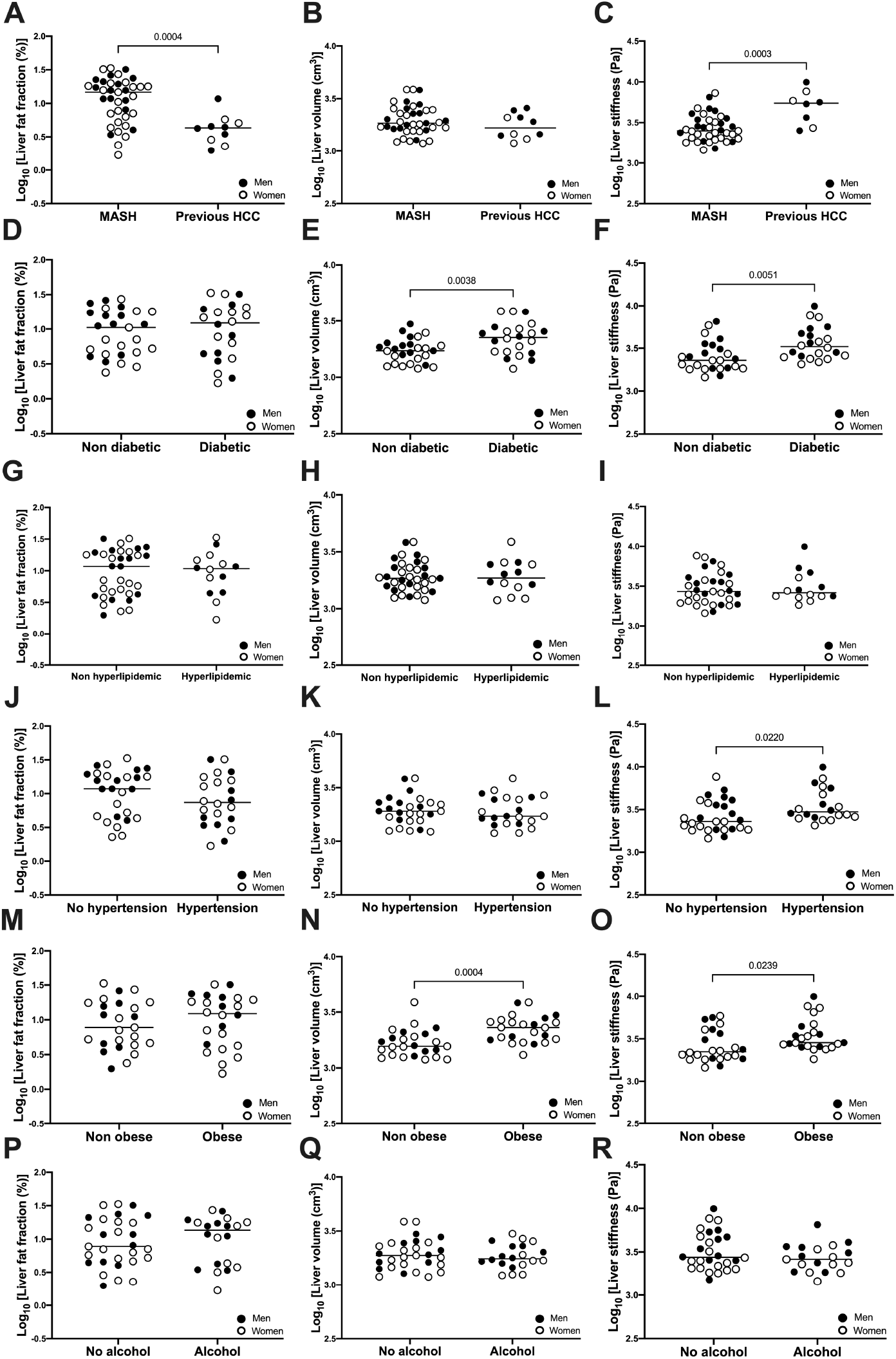
MRE/MRI measures of Liver fat fraction, volume and stiffness in patients with MASH with or without previous history of HCC (A-C), diabetes (D-F), hyperlipidemia (G-I), hypertension (J-L), obesity (M-O) and alcohol consumption (P-R). Data is presented as the distribution around the mean. Statistical significance was evaluated by un-paired t test. For liver fat fraction and liver volume, n=28 women (open circles) and n=21 men (closed circles). For liver stiffness, n=27 women (open circles) and n=20 men (closed circles).

**Supplementary Figure 3.**
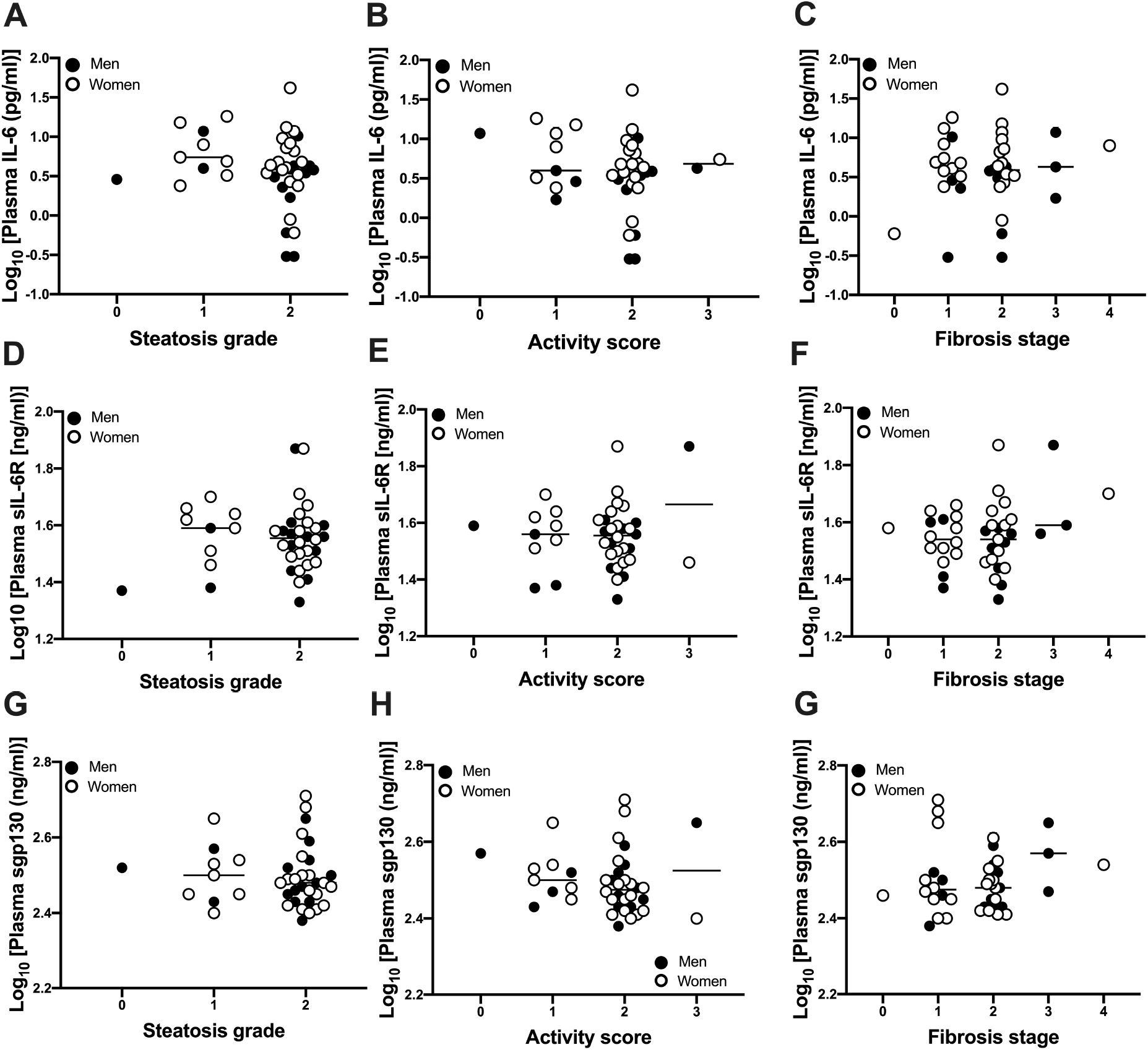
Plasma IL-6 (A-C), sIL-6R (D-F) and sgp130 (G-I) levels in patients with MASH compared along steatosis grade, activity score and fibrosis stage. Patient livers with steatosis grade G0 (n=1), G1 (n=9), G2 (n=37), activity score A0 (n=1 A1 (n=9), A2 (n=37), A3 (n=2), A4 (n=5) and fibrosis stage F0 (n=1), F1 (n=14), F2 (n=21), F3 (n=3), F4 (n=1). Data is presented as the distribution around the mean. Analysis performed by One-way Anova with multiple comparisons. Analysis was adjusted for age, sex, BMI and diabetes. For steatosis grade, activity score and fibrosis stage N=25 women (open circles) and N=15 men (closed circles).

**Supplementary Figure 4.**
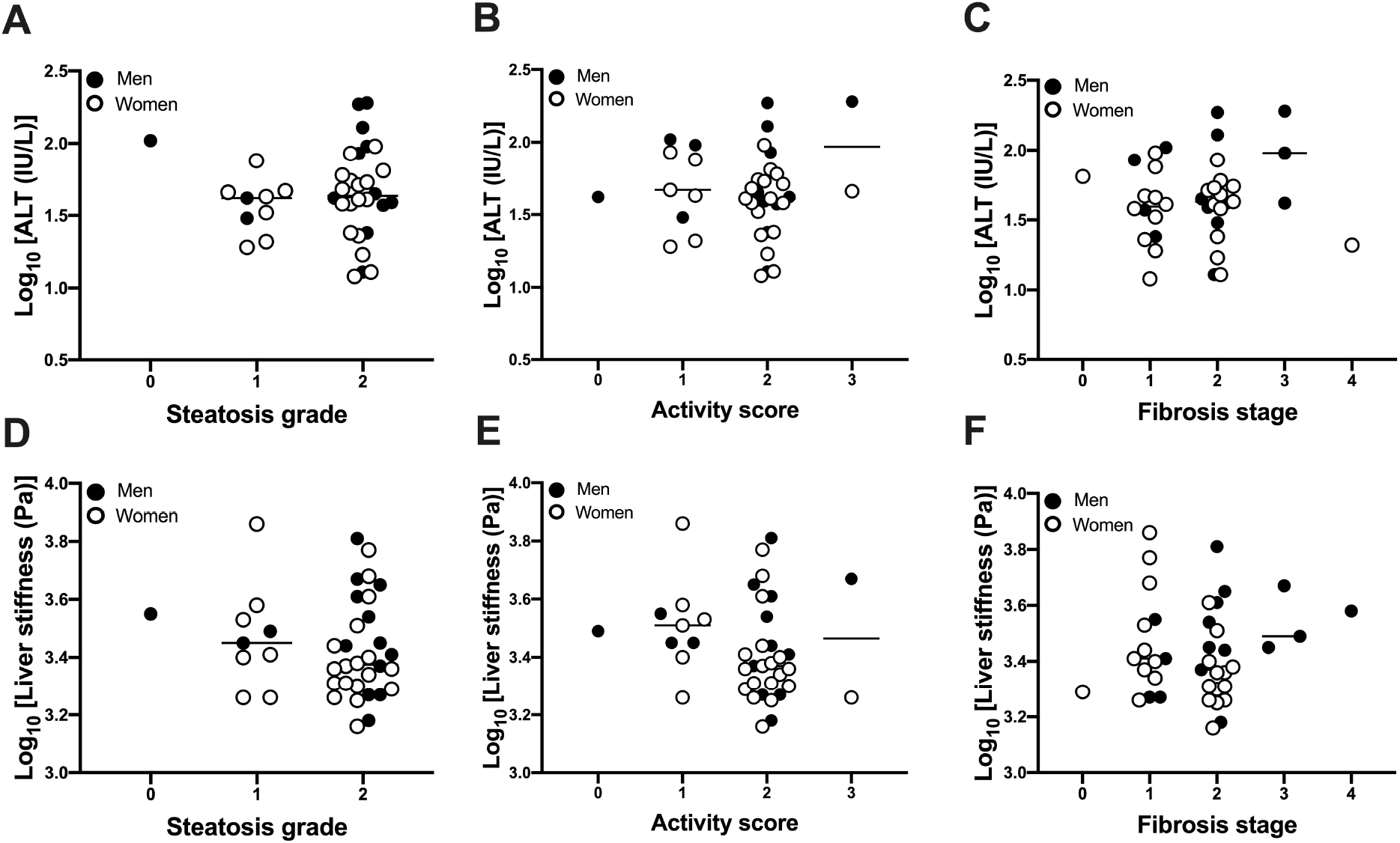
Plasma ALT (A-C) and liver stiffness (D-F) levels measured by MRI/MRE in patients with MASH compared along steatosis grade, activity score and fibrosis stage. Patient livers with steatosis grade G0 (n=1), G1 (n=9), G2 (n=37), activity score A0 (n=1 A1 (n=9), A2 (n=37), A3 (n=2), A4 (n=5) and fibrosis stage F0 (n=1), F1 (n=14), F2 (n=21), F3 (n=3), F4 (n=1). Data is presented as the distribution around the mean. Analysis performed by One-way Anova with multiple comparisons. Analysis was adjusted for age, sex, BMI and diabetes. For steatosis grade, activity score and fibrosis stage N=24 women (open circles) and N=14 men (closed circles).

**Supplementary Figure 5.**
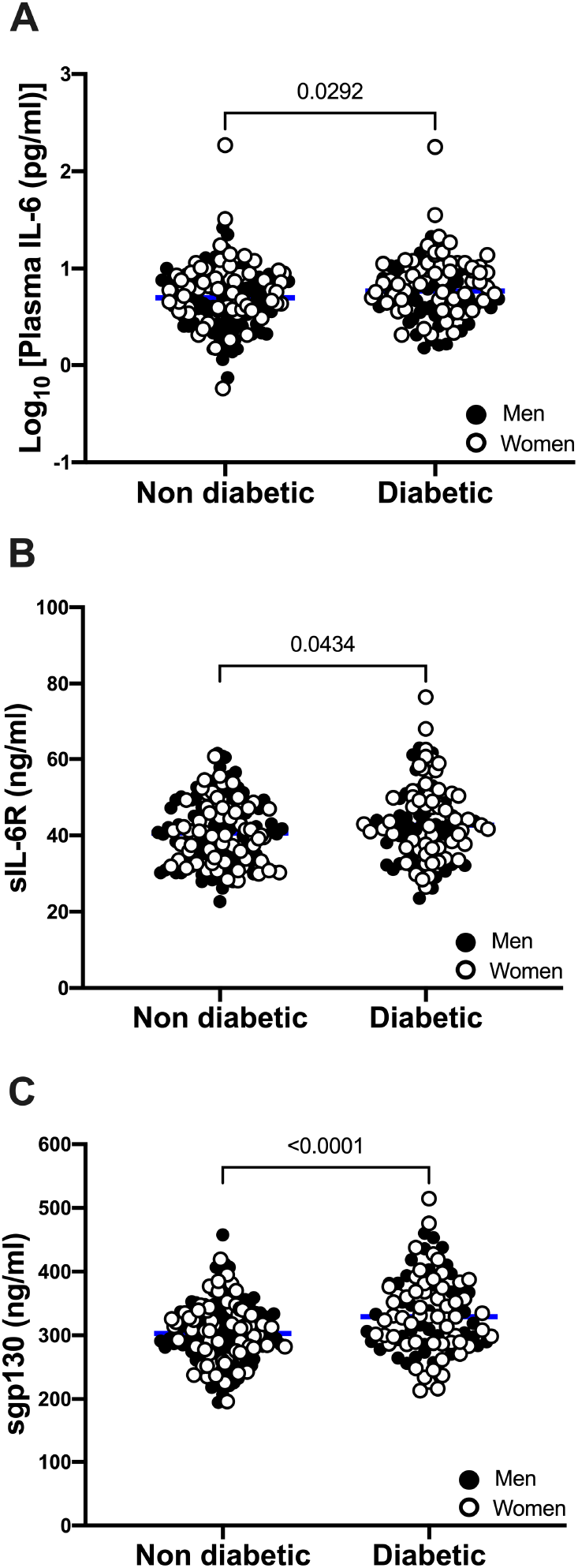
Plasma levels of IL-6 (A), sIL-6R (B) and sgp130 (C) in patients with morbid obesity stratified by diagnosis of diabetes. Data is presented as the distribution around the mean. Statistical significance was evaluated by unpaired t test.

**Supplementary Figure 6.**
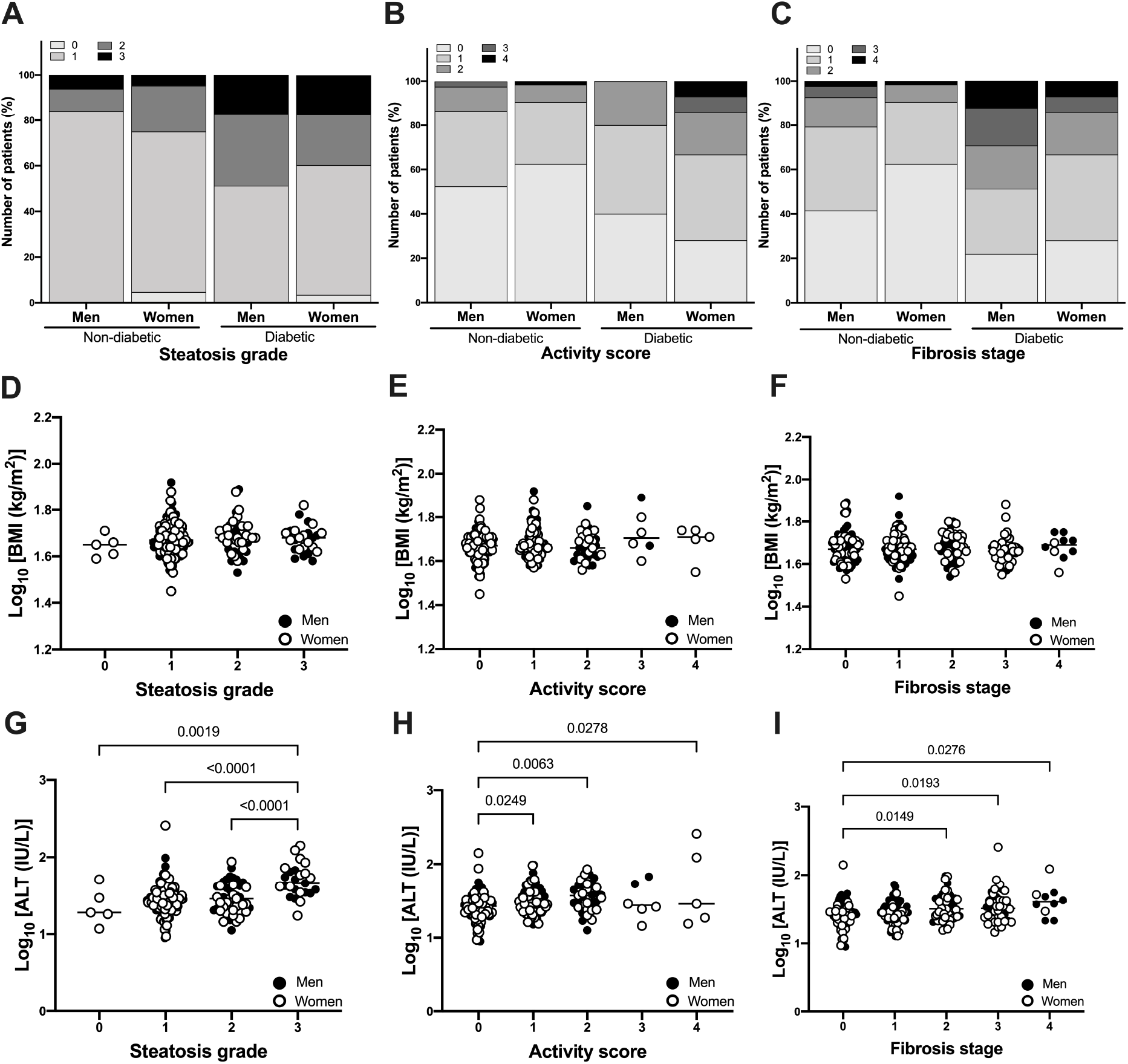
Liver steatosis grade, activity score and fibrosis stage in patients with morbid obesity with or without diabetes (A, D, G), BMI (B, E, H), and plasma ALT concentrations (C, F, I) in relative to various levels of steatosis grade G0 (n=5), G1 (n=168), G2 (n=47), G3 (n=25), activity score A0 (n=115), A1 (n=84), A2 (n=33), A3 (n=6), A4 (n=5) and fibrosis stage F0 (n=73), F1 (n=73), F2 (n=48), F3 (n=41), F4 (n=10) determined by histology. Data is presented as the distribution around the mean. Analysis performed by One-way Anova with multiple comparisons. Analysis was adjusted for age, sex, BMI and diabetes. For activity score N=121 women (open circles) and N=122 men (closed circles).

**SUPPLEMENTARY TABLE 1.**
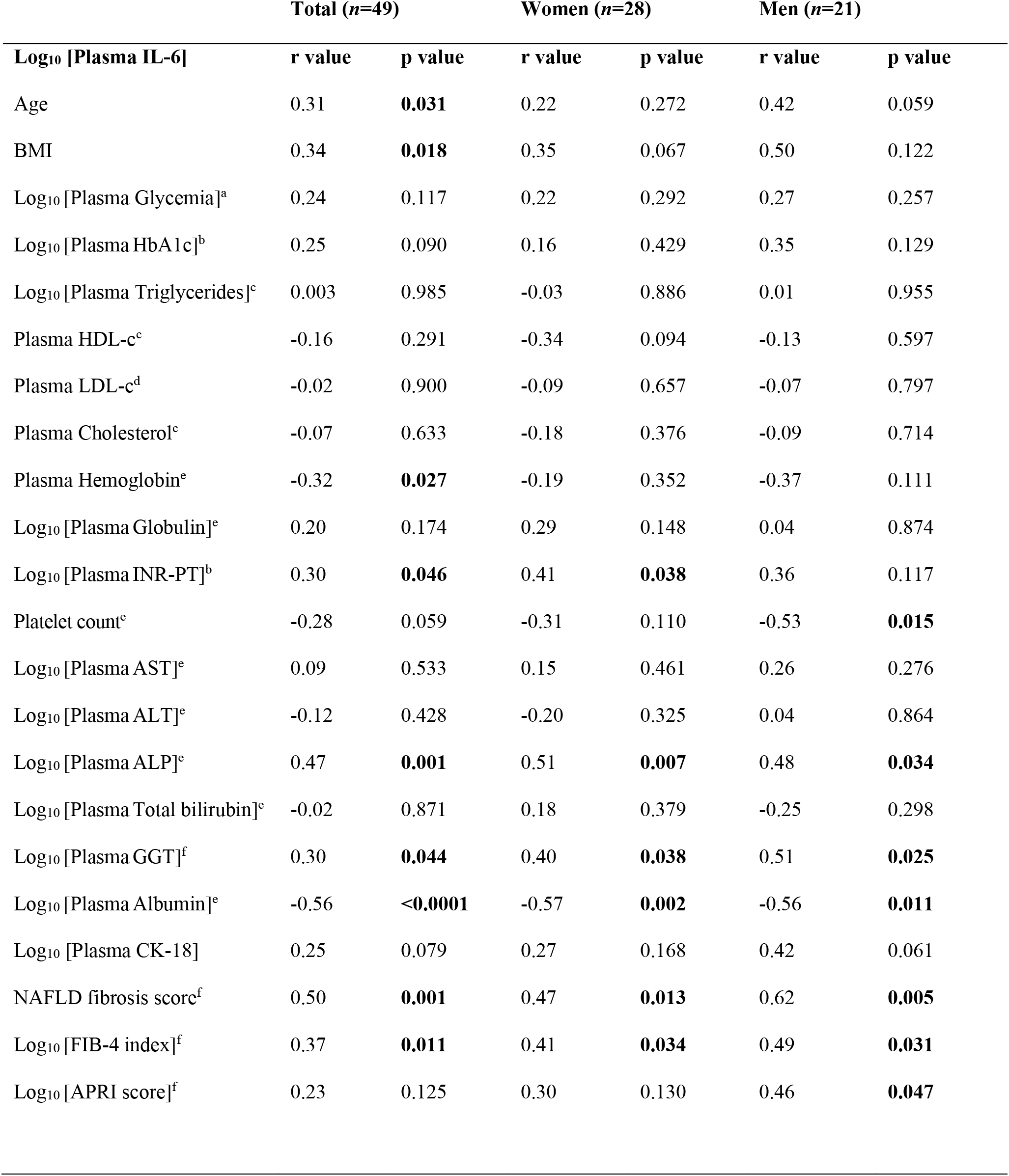
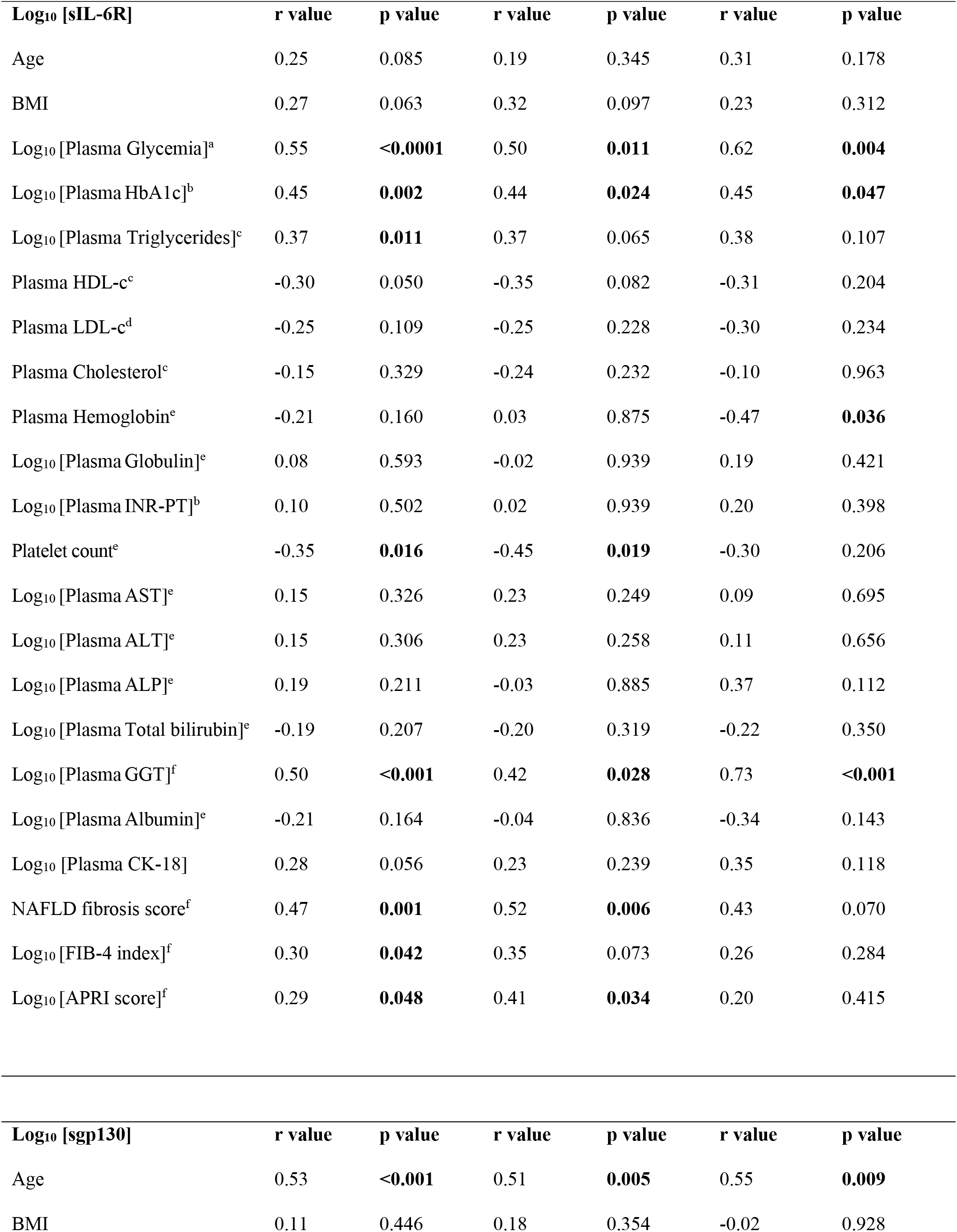
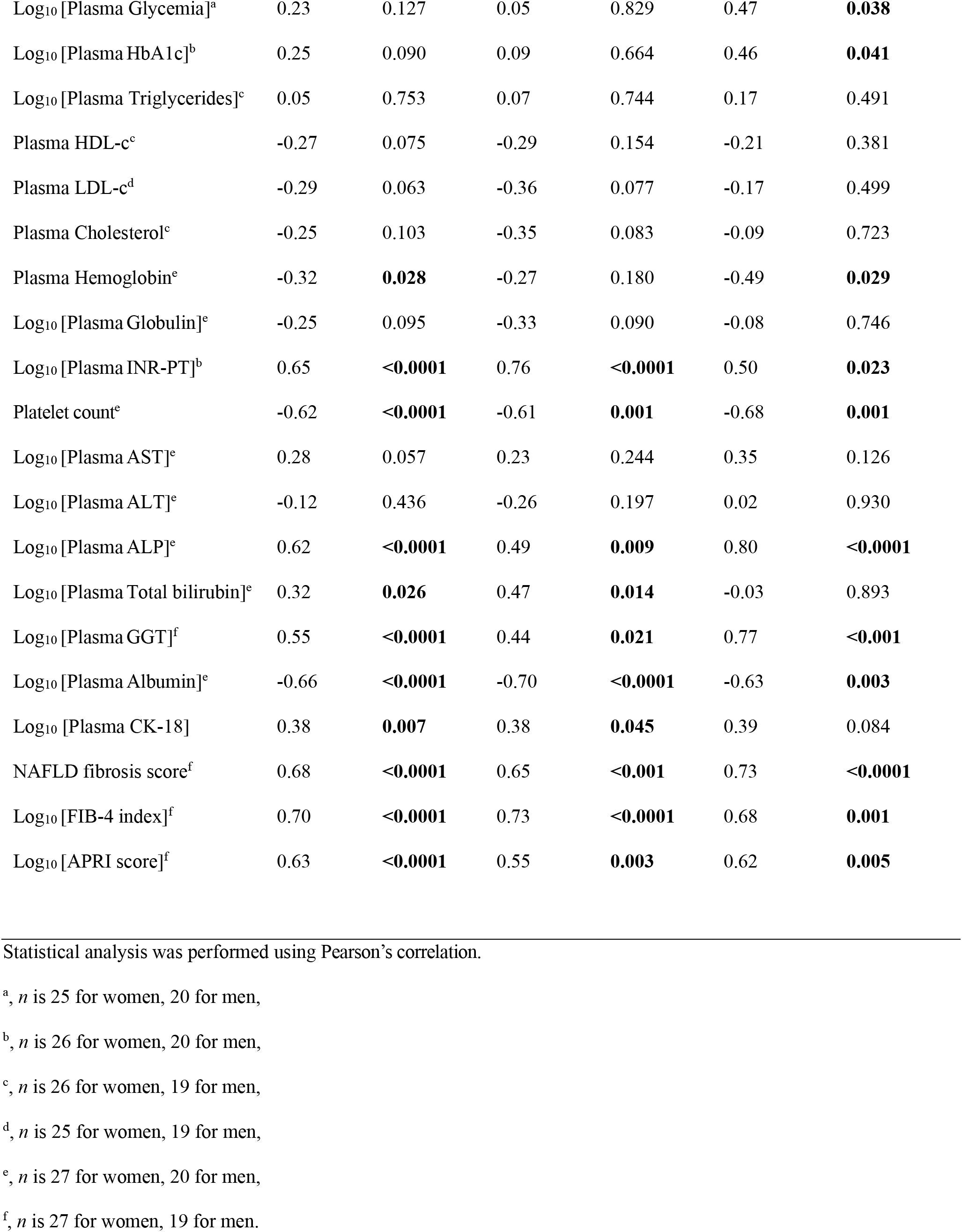
Correlations between Log_10_ plasma IL-6, sIL-6R, sgp130 and common indicators of liver stiffness/fibrosis in patients with MASH.

## REFERENCES

1. Younossi ZM, Koenig AB, Abdelatif D, Fazel Y, Henry L, and Wymer M. Global epidemiology of nonalcoholic fatty liver disease-Meta-analytic assessment of prevalence, incidence, and outcomes. Hepatology. 2016;64(1):73–84.

2. Zhou F, Zhou J, Wang W, Zhang XJ, Ji YX, Zhang P, et al. Unexpected Rapid Increase in the Burden of NAFLD in China From 2008 to 2018: A Systematic Review and Meta-Analysis. Hepatology. 2019;70(4):1119–33.

3. Li J, Zou B, Yeo YH, Feng Y, Xie X, Lee DH, et al. Prevalence, incidence, and outcome of non-alcoholic fatty liver disease in Asia, 1999-2019: a systematic review and meta-analysis. Lancet Gastroenterol Hepatol. 2019;4(5):389–98.

4. Paik JM, Henry L, De Avila L, Younossi E, Racila A, and Younossi ZM. Mortality Related to Nonalcoholic Fatty Liver Disease Is Increasing in the United States. Hepatol Commun. 2019;3(11):1459–71.

5. Orci LA, Sanduzzi-Zamparelli M, Caballol B, Sapena V, Colucci N, Torres F, et al. Incidence of Hepatocellular Carcinoma in Patients With Nonalcoholic Fatty Liver Disease: A Systematic Review, Meta-analysis, and Meta-regression. Clin Gastroenterol Hepatol. 2022;20(2):283–92 e10.

6. Swain MG, Ramji A, Patel K, Sebastiani G, Shaheen AA, Tam E, et al. Burden of nonalcoholic fatty liver disease in Canada, 2019-2030: a modelling study. CMAJ Open. 2020;8(2):E429–E36.

7. Simon TG, Roelstraete B, Khalili H, Hagstrom H, and Ludvigsson JF. Mortality in biopsy-confirmed nonalcoholic fatty liver disease: results from a nationwide cohort. Gut. 2021;70(7):1375–82.

8. Younossi ZM, Stepanova M, Rafiq N, Henry L, Loomba R, Makhlouf H, et al. Nonalcoholic steatofibrosis independently predicts mortality in nonalcoholic fatty liver disease. Hepatol Commun. 2017;1(5):421–8.

9. Eslam M, Newsome PN, Sarin SK, Anstee QM, Targher G, Romero-Gomez M, et al. A new definition for metabolic dysfunction-associated fatty liver disease: An international expert consensus statement. J Hepatol. 2020;73(1):202–9.

10. Fracanzani AL, Valenti L, Bugianesi E, Andreoletti M, Colli A, Vanni E, et al. Risk of severe liver disease in nonalcoholic fatty liver disease with normal aminotransferase levels: a role for insulin resistance and diabetes. Hepatology. 2008;48(3):792–8.

11. Uhanova JM, G; Premji, K; Chandok N.;. Serial liver transaminases have no prognostic value in non-alcoholic fatty liver disease. Canadian Liver Journal. 2019;2(1):19–22.

12. Portillo-Sanchez P, Bril F, Maximos M, Lomonaco R, Biernacki D, Orsak B, et al. High Prevalence of Nonalcoholic Fatty Liver Disease in Patients With Type 2 Diabetes Mellitus and Normal Plasma Aminotransferase Levels. J Clin Endocrinol Metab. 2015;100(6):2231–8.

13. Ma X, Liu S, Zhang J, Dong M, Wang Y, Wang M, et al. Proportion of NAFLD patients with normal ALT value in overall NAFLD patients: a systematic review and meta-analysis. BMC Gastroenterol. 2020;20(1):10.

14. Bastard JP, Jardel C, Bruckert E, Blondy P, Capeau J, Laville M, et al. Elevated levels of interleukin 6 are reduced in serum and subcutaneous adipose tissue of obese women after weight loss. J Clin Endocrinol Metab. 2000;85(9):3338–42.

15. Yeste D, Vendrell J, Tomasini R, Broch M, Gussinye M, Megia A, et al. Interleukin-6 in obese children and adolescents with and without glucose intolerance. Diabetes Care. 2007;30(7):1892–4.

16. Charles BA, Doumatey A, Huang H, Zhou J, Chen G, Shriner D, et al. The roles of IL-6, IL-10, and IL-1RA in obesity and insulin resistance in African-Americans. J Clin Endocrinol Metab. 2011;96(12):E2018–22.

17. Weiss TW, Arnesen H, and Seljeflot I. Components of the interleukin-6 transsignalling system are associated with the metabolic syndrome, endothelial dysfunction and arterial stiffness. Metabolism. 2013;62(7):1008–13.

18. Chen H, Zhang X, Liao N, and Wen F. Increased levels of IL-6, sIL-6R, and sgp130 in the aqueous humor and serum of patients with diabetic retinopathy. Mol Vis. 2016;22:1005–14.

19. Wieckowska A, Papouchado BG, Li Z, Lopez R, Zein NN, and Feldstein AE. Increased hepatic and circulating interleukin-6 levels in human nonalcoholic steatohepatitis. Am J Gastroenterol. 2008;103(6):1372–9.

20. Mohamed KA, Mohamed EE, Ahmed DM, Sayed MA, and Hussien AR. A study of Interleukin 6 as a predictive biomarker for development of nonalcholic steatohepatitis in patients with Nonalcholic fatty liver disease. QJM: An International Journal of Medicine. 2020;113(Supplement_1).

21. Labenz C, Toenges G, Huber Y, Nagel M, Marquardt JU, Schattenberg JM, et al. Raised serum Interleukin-6 identifies patients with liver cirrhosis at high risk for overt hepatic encephalopathy. Aliment Pharmacol Ther. 2019;50(10):1112–9.

22. Porta C, De Amici M, Quaglini S, Paglino C, Tagliani F, Boncimino A, et al. Circulating interleukin-6 as a tumor marker for hepatocellular carcinoma. Ann Oncol. 2008;19(2):353–8.

23. Schmidt-Arras D, and Rose-John S. IL-6 pathway in the liver: From physiopathology to therapy. J Hepatol. 2016;64(6):1403–15.

24. The human protein atlas. https://www.proteinatlas.org/search/IL-6R.

25. Uhlen M, Fagerberg L, Hallstrom BM, Lindskog C, Oksvold P, Mardinoglu A, et al. Proteomics. Tissue-based map of the human proteome. Science. 2015;347(6220):1260419.

26. Nikolajuk A, Kowalska I, Karczewska-Kupczewska M, Adamska A, Otziomek E, Wolczynski S, et al. Serum soluble glycoprotein 130 concentration is inversely related to insulin sensitivity in women with polycystic ovary syndrome. Diabetes. 2010;59(4):1026–9.

27. Shriki A, Lanton T, Sonnenblick A, Levkovitch-Siany O, Eidelshtein D, Abramovitch R, et al. Multiple Roles of IL6 in Hepatic Injury, Steatosis, and Senescence Aggregate to Suppress Tumorigenesis. Cancer Res. 2021;81(18):4766–77.

28. Lemmers A, Gustot T, Durnez A, Evrard S, Moreno C, Quertinmont E, et al. An inhibitor of interleukin-6 trans-signalling, sgp130, contributes to impaired acute phase response in human chronic liver disease. Clin Exp Immunol. 2009;156(3):518–27.

29. Reference values for clinical samples, The University of Montreal Hospital Research Centre (CRCHUM). https://www.chumontreal.qc.ca/sites/default/files/inline-files/180710_Valeurs_reference_Biochimie-CHUM_2018-05-08.pdf.

30. Nikolaus S, Waetzig GH, Butzin S, Ziolkiewicz M, Al-Massad N, Thieme F, et al. Evaluation of interleukin-6 and its soluble receptor components sIL-6R and sgp130 as markers of inflammation in inflammatory bowel diseases. Int J Colorectal Dis. 2018;33(7):927–36.

31. Padberg F, Feneberg W, Schmidt S, Schwarz MJ, Körschenhausen D, Greenberg BD, et al. CSF and serum levels of soluble interleukin-6 receptors (sIL-6R and sgp130), but not of interleukin-6 are altered in multiple sclerosis. J Neuroimmunol. 1999;99(2):218–23.

32. Tang A, Cloutier G, Szeverenyi NM, and Sirlin CB. Ultrasound Elastography and MR Elastography for Assessing Liver Fibrosis: Part 1, Principles and Techniques. American Journal of Roentgenology. 2015;205(1):22–32.

33. Costa-Silva L, Ferolla SM, Lima AS, Vidigal PVT, and Ferrari TCA. MR elastography is effective for the non-invasive evaluation of fibrosis and necroinflammatory activity in patients with nonalcoholic fatty liver disease. Eur J Radiol. 2018;98:82–9.

34. Chen J, Yin M, Talwalkar JA, Oudry J, Glaser KJ, Smyrk TC, et al. Diagnostic Performance of MR Elastography and Vibration-controlled Transient Elastography in the Detection of Hepatic Fibrosis in Patients with Severe to Morbid Obesity. Radiology. 2017;283(2):418–28.

35. Park CC, Nguyen P, Hernandez C, Bettencourt R, Ramirez K, Fortney L, et al. Magnetic Resonance Elastography vs Transient Elastography in Detection of Fibrosis and Noninvasive Measurement of Steatosis in Patients With Biopsy-Proven Nonalcoholic Fatty Liver Disease. Gastroenterology. 2017;152(3):598–607 e2.

36. Lefebvre T, Wartelle-Bladou C, Wong P, Sebastiani G, Giard JM, Castel H, et al. Prospective comparison of transient, point shear wave, and magnetic resonance elastography for staging liver fibrosis. Eur Radiol. 2019;29(12):6477–88.

37. Zuliani G, Galvani M, Maggio M, Volpato S, Bandinelli S, Corsi AM, et al. Plasma soluble gp130 levels are increased in older subjects with metabolic syndrome. The role of insulin resistance. Atherosclerosis. 2010;213(1):319–24.

38. Bowker N, Shah RL, Sharp SJ, Luan J, Stewart ID, Wheeler E, et al. Meta-analysis investigating the role of interleukin-6 mediated inflammation in type 2 diabetes. EBioMedicine. 2020;61:103062.

39. Skuratovskaia D, Komar A, Vulf M, Quang HV, Shunkin E, Volkova L, et al. IL-6 Reduces Mitochondrial Replication, and IL-6 Receptors Reduce Chronic Inflammation in NAFLD and Type 2 Diabetes. Int J Mol Sci. 2021;22(4).

40. Roy AK, and Chatterjee B. Sexual dimorphism in the liver. Annu Rev Physiol. 1983;45:37–50.

41. Long MT, Pedley A, Massaro JM, Hoffmann U, Ma J, Loomba R, et al. A simple clinical model predicts incident hepatic steatosis in a community-based cohort: The Framingham Heart Study. Liver Int. 2018;38(8):1495–503.

42. Yang D, Hanna DL, Usher J, LoCoco J, Chaudhari P, Lenz HJ, et al. Impact of sex on the survival of patients with hepatocellular carcinoma: a Surveillance, Epidemiology, and End Results analysis. Cancer. 2014;120(23):3707–16.

43. Bedossa P, Poitou C, Veyrie N, Bouillot JL, Basdevant A, Paradis V, et al. Histopathological algorithm and scoring system for evaluation of liver lesions in morbidly obese patients. Hepatology. 2012;56(5):1751–9.

44. Bergmann J, Muller M, Baumann N, Reichert M, Heneweer C, Bolik J, et al. IL-6 trans-signaling is essential for the development of hepatocellular carcinoma in mice. Hepatology. 2017;65(1):89–103.

45. Fazel Modares N, Polz R, Haghighi F, Lamertz L, Behnke K, Zhuang Y, et al. IL-6 Trans-signaling Controls Liver Regeneration After Partial Hepatectomy. Hepatology. 2019;70(6):2075–91.

46. Hou X, Yin S, Ren R, Liu S, Yong L, Liu Y, et al. Myeloid-Cell-Specific IL-6 Signaling Promotes MicroRNA-223-Enriched Exosome Production to Attenuate NAFLD-Associated Fibrosis. Hepatology. 2021;74(1):116–32.

47. Naeem M, Markus MRP, Mousa M, Schipf S, Dorr M, Steveling A, et al. Associations of liver volume and other markers of hepatic steatosis with all-cause mortality in the general population. Liver Int. 2021.

48. Dong J, Viswanathan S, Adami E, Singh BK, Chothani SP, Ng B, et al. Hepatocyte-specific IL11 cis-signaling drives lipotoxicity and underlies the transition from NAFLD to NASH. Nat Commun. 2021;12(1):66.

49. Kleiner DE, Brunt EM, Van Natta M, Behling C, Contos MJ, Cummings OW, et al. Design and validation of a histological scoring system for nonalcoholic fatty liver disease. Hepatology. 2005;41(6):1313–21.

50. Venkatesh SK, Yin M, and Ehman RL. Magnetic resonance elastography of liver: technique, analysis, and clinical applications. J Magn Reson Imaging. 2013;37(3):544–55.

51. Angulo P, Hui JM, Marchesini G, Bugianesi E, George J, Farrell GC, et al. The NAFLD fibrosis score: a noninvasive system that identifies liver fibrosis in patients with NAFLD. Hepatology. 2007;45(4):846–54.

52. Sterling RK, Lissen E, Clumeck N, Sola R, Correa MC, Montaner J, et al. Development of a simple noninvasive index to predict significant fibrosis in patients with HIV/HCV coinfection. Hepatology. 2006;43(6):1317–25.

53. Wai CT, Greenson JK, Fontana RJ, Kalbfleisch JD, Marrero JA, Conjeevaram HS, et al. A simple noninvasive index can predict both significant fibrosis and cirrhosis in patients with chronic hepatitis C. Hepatology. 2003;38(2):518–26.

54. Brunt EM, Janney CG, Di Bisceglie AM, Neuschwander-Tetri BA, and Bacon BR. Nonalcoholic steatohepatitis: a proposal for grading and staging the histological lesions. Am J Gastroenterol. 1999;94(9):2467–74.

